# Host-Microbe Multiomic Profiling Reveals Age-Dependent COVID-19 Immunopathology

**DOI:** 10.1101/2024.02.11.24301704

**Authors:** Hoang Van Phan, Alexandra Tsitsiklis, Cole P. Maguire, Elias K. Haddad, Patrice M. Becker, Seunghee Kim-Schulze, Brian Lee, Jing Chen, Annmarie Hoch, Harry Pickering, Patrick Van Zalm, Matthew C. Altman, Alison D. Augustine, Carolyn S. Calfee, Steve Bosinger, Charles Cairns, Walter Eckalbar, Leying Guan, Naresh Doni Jayavelu, Steven H. Kleinstein, Florian Krammer, Holden T. Maecker, Al Ozonoff, Bjoern Peters, Nadine Rouphael, IMPACC Network, Ruth R. Montgomery, Elaine Reed, Joanna Schaenman, Hanno Steen, Ofer Levy, Joann Diray-Arce, Charles R. Langelier

## Abstract

Age is a major risk factor for severe coronavirus disease-2019 (COVID-19), yet the mechanisms responsible for this relationship have remained incompletely understood. To address this, we evaluated the impact of aging on host and viral dynamics in a prospective, multicenter cohort of 1,031 patients hospitalized for COVID-19, ranging from 18 to 96 years of age. We performed blood transcriptomics and nasal metatranscriptomics, and measured peripheral blood immune cell populations, inflammatory protein expression, anti-SARS-CoV-2 antibodies, and anti-interferon (IFN) autoantibodies. We found that older age correlated with an increased SARS-CoV-2 viral load at the time of admission, and with delayed viral clearance over 28 days. This contributed to an age-dependent increase in type I IFN gene expression in both the respiratory tract and blood. We also observed age-dependent transcriptional increases in peripheral blood IFN-(, neutrophil degranulation, and Toll like receptor (TLR) signaling pathways, and decreases in T cell receptor (TCR) and B cell receptor signaling pathways. Over time, older adults exhibited a remarkably sustained induction of proinflammatory genes (e.g., CXCL6) and serum chemokines (e.g., CXCL9) compared to younger individuals, highlighting a striking age-dependent impairment in inflammation resolution. Augmented inflammatory signaling also involved the upper airway, where aging was associated with upregulation of TLR, IL17, type I IFN and IL1 pathways, and downregulation TCR and PD-1 signaling pathways. Metatranscriptomics revealed that the oldest adults exhibited disproportionate reactivation of herpes simplex virus and cytomegalovirus in the upper airway following hospitalization. Mass cytometry demonstrated that aging correlated with reduced naïve T and B cell populations, and increased monocytes and exhausted natural killer cells. Transcriptional and protein biomarkers of disease severity markedly differed with age, with the oldest adults exhibiting greater expression of TLR and inflammasome signaling genes, as well as proinflammatory proteins (e.g., IL6, CXCL8), in severe COVID-19 compared to mild/moderate disease. Anti-IFN autoantibody prevalence correlated with both age and disease severity. Taken together, this work profiles both host and microbe in the blood and airway to provide fresh insights into aging-related immune changes in a large cohort of vaccine-naïve COVID-19 patients. We observed age-dependent immune dysregulation at the transcriptional, protein and cellular levels, manifesting in an imbalance of inflammatory responses over the course of hospitalization, and suggesting potential new therapeutic targets.

**One sentence summary:** We observed age-dependent immune dysregulation at the transcriptional, protein and cellular levels, manifesting in an imbalance of inflammatory responses over the course of hospitalization, and suggesting potential new therapeutic targets.

## Introduction

Age is a major risk factor for severe Coronavirus disease 2019 (COVID-19), with older adults experiencing markedly greater rates of acute respiratory distress syndrome (ARDS) and death compared to younger indiviudals^1–3^. Even with vaccination rates above 90%, adults over 75 years of age are 140 times more likely to die if infected with SARS-CoV-2^4^. Despite these striking epidemiological associations, the biological mechanisms underlying the impact of aging on COVID-19 remain incompletely understood.

Observational cohort studies of healthy adults^5,6^ demonstrate that aging leads to baseline increases in plasma concentrations of proinflammatory cytokines^5,7^, several of which (e.g., IL6) are well-known biomarkers of COVID-19 severity, suggesting potential connections between the pathophysiology of human aging and COVID-19^7^. Juxtaposed against this state of aging-associated inflammation are functional impairments in innate and adaptive immune signaling, observed during vaccination of aged individuals^8–11^. Furthermore, recent human *in vitro* data indicates that aging results in impaired production of type I interferons in monocytes and dendritic cells following Toll-like receptor (TLR) ligation, suggesting disrupted innate immunity^12–16^.

Comparative upper respiratory tract transcriptional profiling has demonstrated that mild SARS-CoV-2 infection induces a more robust innate and adaptive immune response in children compared to adults^17,18^. Paradoxically, amongst adults hospitalized for COVID-19, a more robust immune response underlies the pathogenesis of severe disease, suggesting more complicated relationships between aging and host defense for older individuals. Adding further complexity, and highlighting the need for additional investigation, is the association between increased age, development of anti-interferon autoantibodies (autoAbs), and disease severity^2,19,20^.

The pathophysiology of COVID-19 involves a dynamic relationship between SARS-CoV-2 and the host immune response^21,22^, yet studies of COVID-19 and aging have assessed each independently. Furthermore, heterogeneity in human physiology necessitates a large sample size to optimally study aging and host immunity. To address these gaps, we leverage data from 1,031 adults hospitalized for COVID-19 enrolled in the IMPACC (IMmunoPhenotyping Assessment in a COVID-19 Cohort) cohort^2,23^, and perform a multiomic, host/microbe systems immunoprofiling study of aging.

From 2,523 longitudinally collected blood and nasal swab samples, we investigate the impact of aging on SARS-CoV-2 viral load, SARS-CoV-2 antibody (Ab) levels, host gene expression, inflammatory protein expression, immune cell populations, and the nasal microbiome. Our study leverages a robust multicenter cohort to gain new insights into aging and immunity by integrating host and microbe data. This work builds on landmark clinical studies demonstrating that age is a major risk factor for COVID-19 severity^1–3^, assesses the generalizability of early translational studies that included smaller numbers of study participants^7,12,24,25^, and generates fresh insights into age-dependent COVID-19 pathophysiology that could assist in developing age-specific therapeutic interventions and biomarkers of disease severity.

## Results

### Study cohort

We analyzed blood and nasal swab specimens from 1,031 adults with COVID-19 enrolled in the IMPACC cohort from 20 hospitals across the United States^2,23^ (Fig. 1, Supp. Table 1). Participants were grouped into quintiles based on age for analyses (18-46, 47-54, 55-62, 63-70, and 71-96 years old), ranging from 187 to 223 participants (median 206 participants) per age group (Fig. 2a). We analyzed age distributions across five previously defined COVID-19 disease trajectory groups^2^, ranging from mild disease with brief length of hospital stay (TG1) to severe disease and death (TG5). We found that advanced age was significantly associated with disease trajectory group (Fig. 2b) and mortality (Fig. 2c). To investigate host immunologic and microbial features associated with age, we employed a wide range of assays at baseline (within 72 hours of hospital admission) and longitudinally post-hospital admission (Fig. 1, Methods). These included transcriptional profiling of PBMCs and nasal swabs, soluble serum immune protein profiling, whole blood mass cytometry (CyTOF), nasal metatranscriptomics, SARS-CoV-2 IgG assays, and anti-IFN-α autoAb measurements.

**Fig. 1:**
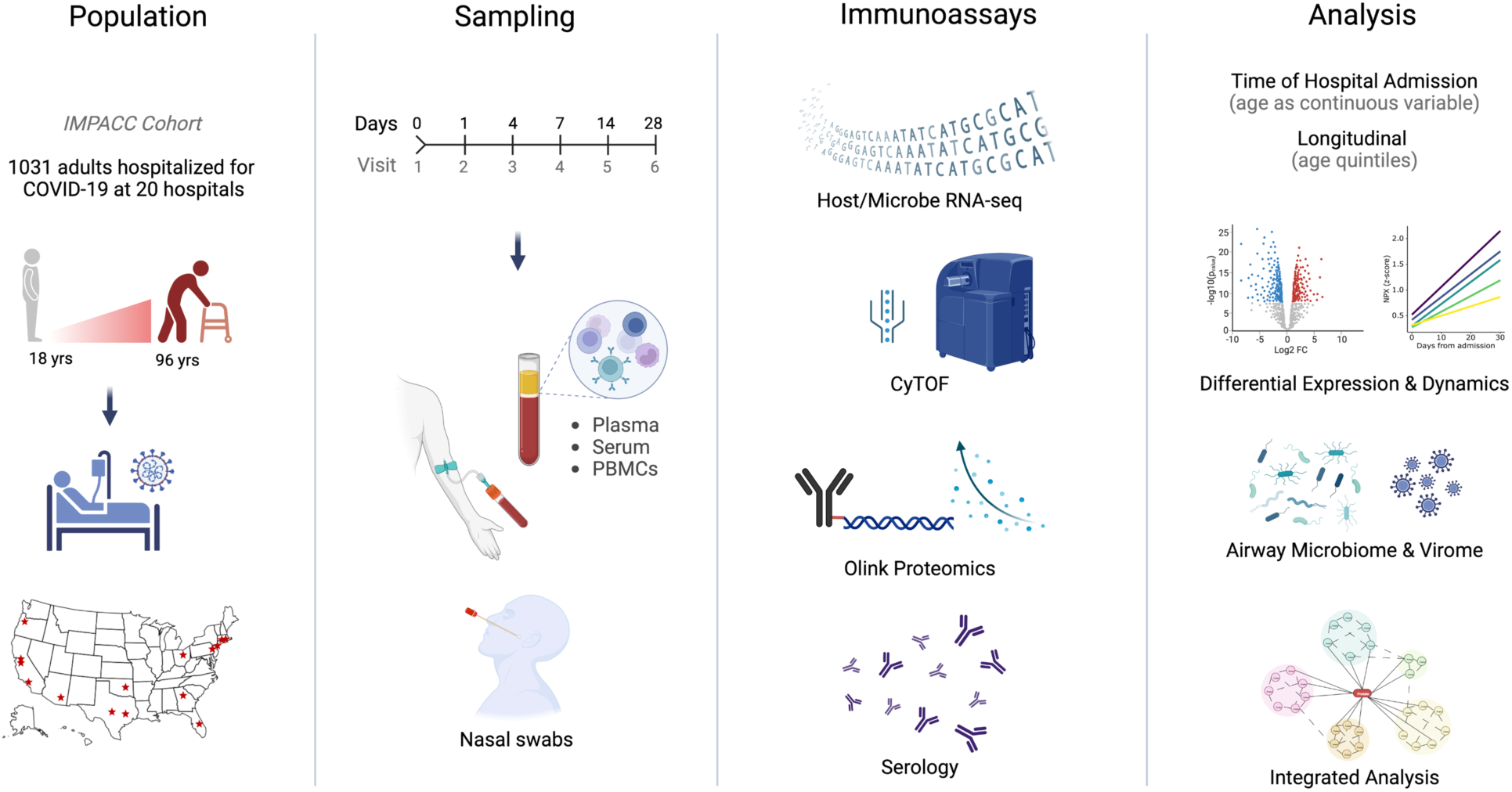
Graphical study overview. This study evaluated 1,031 COVID-19 patients between the ages of 18 and 96 enrolled in the IMPACC cohort at 20 hospitals across the United States. Blood (PBMCs, plasma and serum) and nasal swab samples were collected at up to 6 visits over 28 days and processed for RNA sequencing, proteomics, mass cytometry, and serology.

**Fig. 2:**
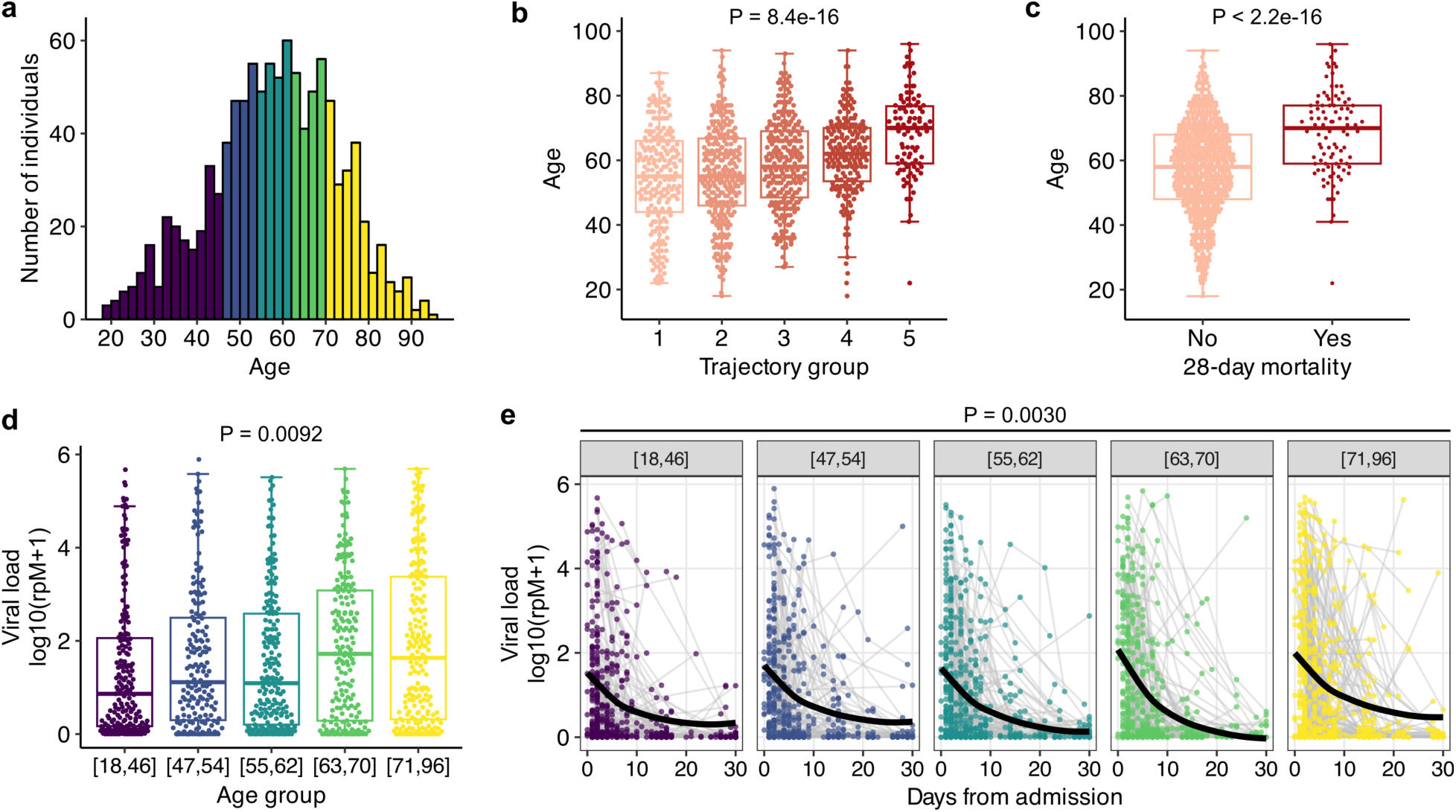
Older adults have more severe COVID-19 and higher SARS-CoV-2 viral loads. (a) Age distribution of the participant cohort. (b, c) Box plot showing the relationship between patients’ age and (b) trajectory group severity or (c) mortality. (d) Nasal swab SARS-CoV-2 viral load at Visit 1 (reads per million (rpM) measured by metatranscriptomics) in each age group. In (b-d), P-values were calculated using Kruskal-Wallis test. (e) Nasal swab SARS-CoV-2 viral load over time in each age group. P-value was calculated with generalized additive mixed effects modeling.

### Aging is associated with higher SARS-CoV-2 viral load, impaired viral clearance, and lower SARS-CoV-2 antibody levels

Older adults had a significantly higher SARS-CoV-2 viral load at Visit 1, measured in reads per million (rpM) from nasal swab RNA sequencing (P = 0.0011, Fig. 2d), a measurement that highly correlated with qPCR cycle threshold (P < 2.2e-16, Supp. Fig. 1). This age-related increase in viral load was not explained by differences in time from symptom onset (Supp. Fig. 2). Longitudinal analysis also revealed significant differences in viral load dynamics, with the oldest adults demonstrating reduced viral clearance compared to the youngest adults (P = 0.0024, Fig. 2e). We also assessed anti-SARS-CoV-2 receptor binding domain (RBD) IgG levels across the five age groups, and found that the oldest adults had lower levels at Visit 1 (Supp. Fig. 3a) and lower Ab production over time (Supp. Fig. 3b).

### Age-dependent differences in immune cell populations

We quantified differences in proportions of immune cell populations in the peripheral blood by mass cytometry (CyTOF) to assess whether aging altered cell frequencies^23^. Using a panel of 43 Abs designed to identify cell lineages in whole blood samples from Visit 1, we found 21 cell types (Fig. 3a) significantly associated with participant age (adjusted P < 0.05, Fig. 3b). Increased age correlated with higher proportions of circulating classical monocytes (CD14+ CD16-), non-classical monocytes (CD14-CD16+), and intermediate monocytes (CD14+ CD16+). Terminally differentiated/exhausted natural killer (NK) cell (CD56^low^ CD16^hi^ CD57^hi^) proportions also increased with age, as did activated CD4+ T cells and central memory (CM) CD8+ T cells (Fig. 3b, c). In contrast, older adults had lower proportions of naïve CD8+ T cells, naïve B cells, gamma-delta (γδ) T cells, and plasmablasts (Fig. 3c). Finally, we found that the age-related differences in cell type frequencies were not affected by SARS-CoV-2 viral load (Supp. Fig. 4).

**Fig. 3:**
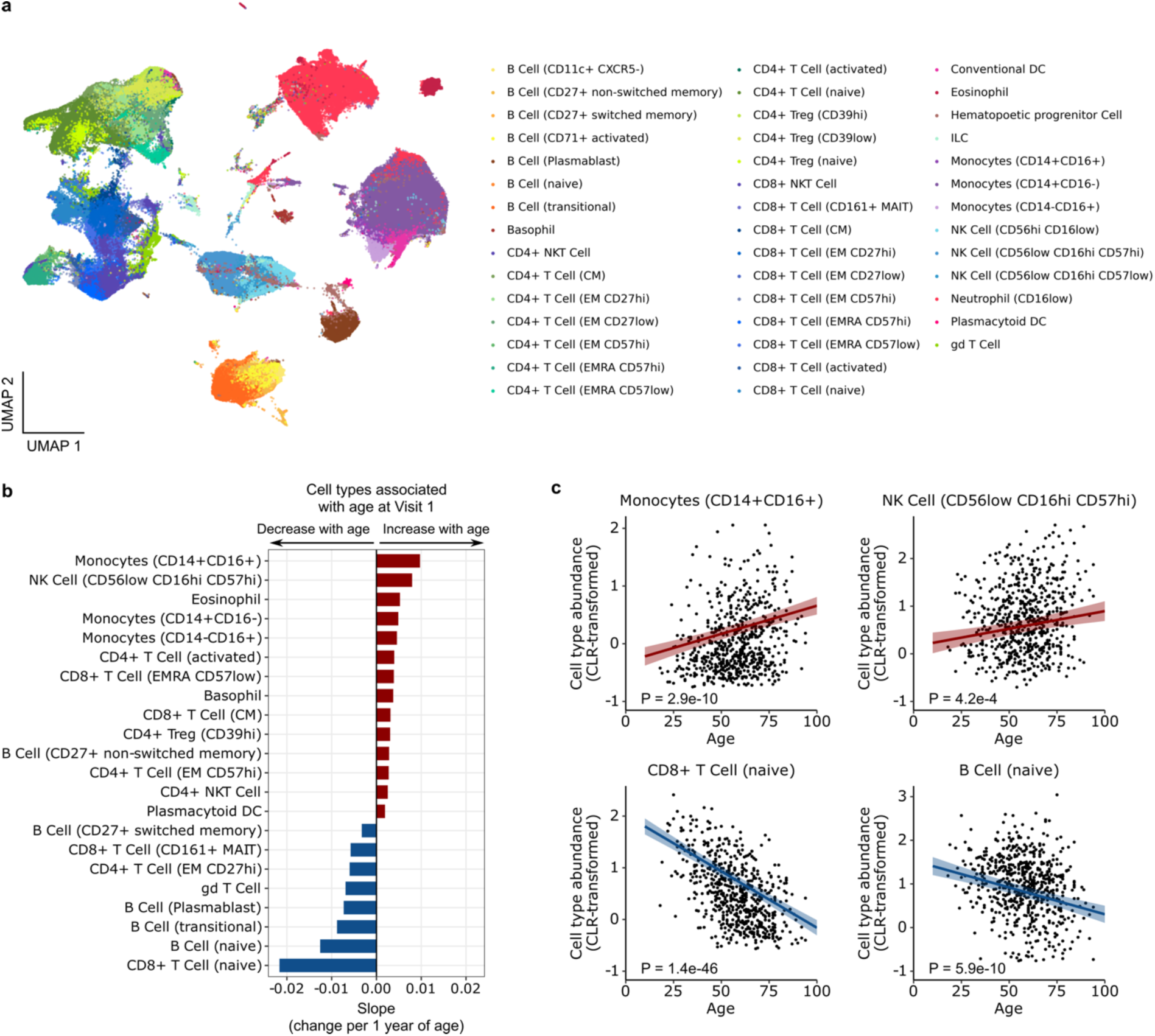
Aging alters immune cell populations during COVID-19. (a) Uniform Manifold Approximation and Projection (UMAP) plot highlighting blood cell types analyzed by CyTOF. (b) Bar plot depicting blood cell types that are upregulated (red) or downregulated (blue) with age at Visit 1. “gd T cell” stands for γδ T cell. (c) Scatter plots depict centered log ratio (CLR) transformed proportions of CD14+CD16+ monocytes and naïve CD8 T cells as a function of age. P values were calculated using linear modeling with Benjamini-Hochberg correction.

### Age-dependent changes in PBMC gene expression at the time of hospitalization

Next, we performed PBMC transcriptional profiling and identified 3,763 genes significantly associated with age at baseline (Visit 1), controlling for sex and disease severity (see Methods) (adjusted P < 0.05, Fig. 4a). Gene set enrichment analysis (GSEA) revealed upregulation of several innate immune-related pathways in older participants, including IFN-α/β and TLR cascades, as well as IFN-γ signaling (Fig. 4b). In contrast, several adaptive immunity-related pathways were downregulated in older individuals, such as B cell receptor (BCR) signaling, T cell receptor (TCR) signaling, and PD1 signaling.

**Fig. 4:**
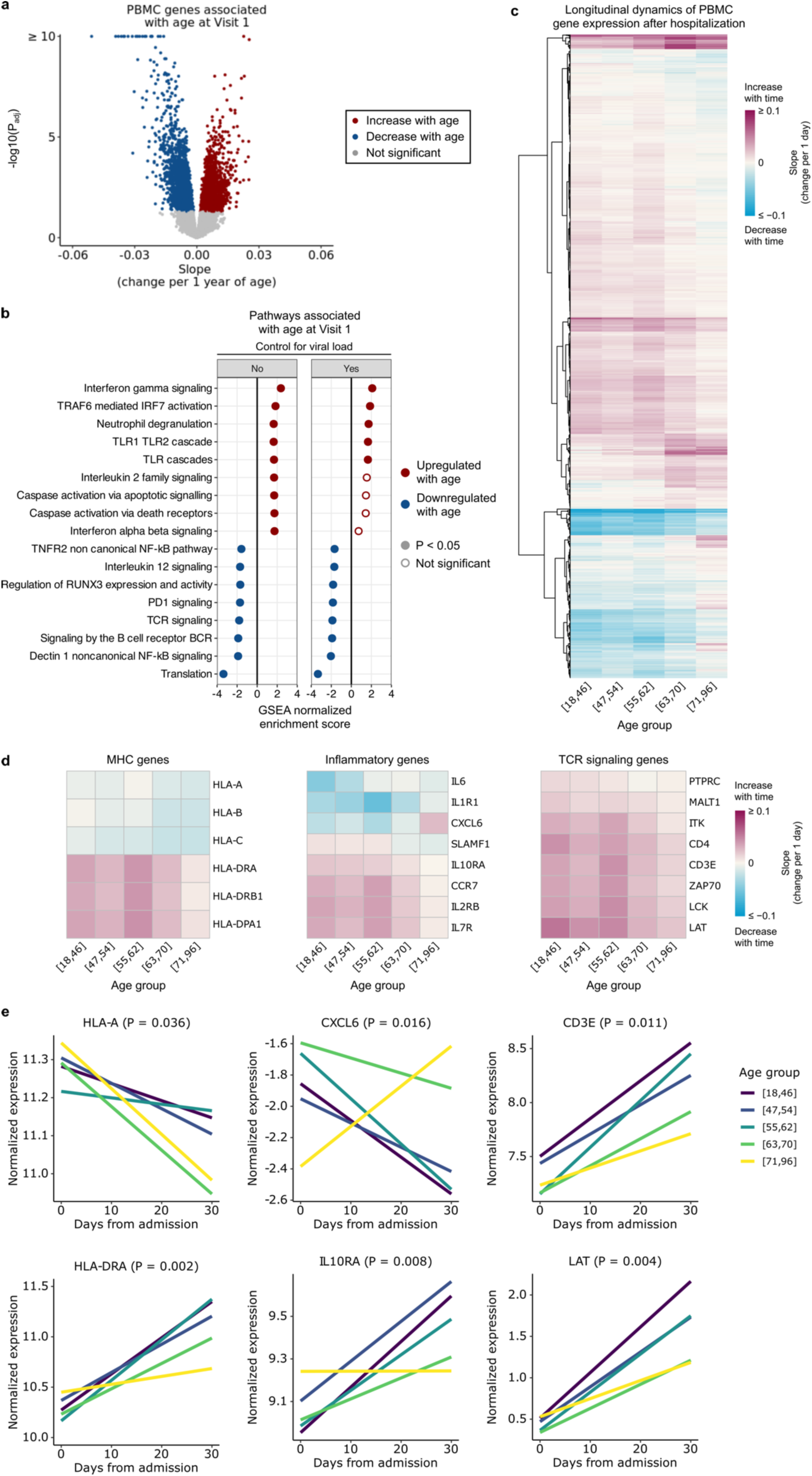
Aging leads to changes in PBMC gene expression during COVID-19. (a) Volcano plot highlighting genes associated with age at Visit 1 in PBMC RNA-seq data. (b) Plot demonstrating the normalized enrichment score of select Reactome pathways associated with age at Visit 1, with and without controlling for viral load, in PBMC samples. (Full results are tabulated in Supp. Data 1 and 2.) P values in (a, b) were calculated with limma’s linear model and Benjamini-Hochberg correction. (c) Heatmap representing the temporal slopes (i.e., change in gene expression per 1 day) of 2,812 genes that differ longitudinally between the 5 age groups (adjusted P < 0.05). (d) Heatmaps representing the temporal slopes of select MHC, inflammatory, and TCR signaling genes from (c). (e) Plots demonstrating the temporal dynamics of 6 example genes from (g). P values were calculated using linear mixed effects modeling and Benjamini-Hochberg correction. (Full temporal dynamics plots with confidence intervals are provided in Supp. Fig. 6.)

### Increased activation of type I interferon signaling in older adults

Prior studies have demonstrated a direct relationship between SARS-CoV-2 viral load and IFN-stimulated gene (ISG) expression^17,26^. We therefore hypothesized that the strong positive correlation between age and viral load (Fig. 2e) might contribute to the upregulation of innate immunity genes and pathways that we observed in older adults. To test this hypothesis, we repeated the differential expression and GSEA analyses while controlling for SARS-CoV-2 viral load. Age-related increases in IFN-γ, TLR signaling, and neutrophil degranulation remained significant. IFN-α/β, IL2, and caspase activation signaling, however, lost statistical significance, suggesting that the stronger activation of these pathways in older patients was due to the age-related increase in viral load.

To assess whether our observations were specific for COVID-19 or reflected general effects of aging, we compared our GSEA results against public data from 14,983 healthy adults across the age spectrum^5^. While we observed age-related upregulation of some pathways (e.g., IFN-γ and TLR signaling) in both our data and the healthy controls (Supp. Fig. 5a), other pathways were uniquely upregulated in the context of COVID-19 (e.g., caspase activation, TRAF6-mediated IRF7 activation) (Supp. Data 3). Similarly, age-related downregulation of TCR and BCR signaling, as well as several other pathways, was unique to COVID-19 (Supp. Fig. 5b).

### Age-dependent differences in the longitudinal dynamics of PBMC gene expression

We next performed a longitudinal analysis of PBMC transcriptomics data over 28 days following hospital admission to identify genes that exhibited age-dependent differences in temporal dynamics, while controlling for participants’ sex and severity trajectory group. Using linear mixed effects modeling, we identified 2,737 genes that had different longitudinal dynamics across age quintiles (Fig. 4c, Supp. Data 5). Several groups of genes demonstrated marked differences in expression dynamics. For example, the expression of MHC class II genes (e.g., *HLA-DRA*) increased over time post-hospitalization in all age groups, but the rate of increase was greater in younger participants (Fig. 4d, e, Supp. Fig. 6). In contrast, the expression of MHC class I genes (e.g., *HLA-A*) decreased over time across all five age groups, but the rate of decrease was greater in older participants. TCR signaling genes (e.g., *CD3E*, *LAT*) were globally upregulated over time, however their induction was notably attenuated in the oldest versus youngest age quintiles.

The longitudinal dynamics of several canonical inflammatory genes also differed between age groups. For instance, the expression of *CXCL6* increased over the course of hospitalization in the oldest age group, while in younger participants its expression decreased markedly. In contrast, expression of the anti-inflammatory gene *IL10RA* in the youngest participants increased over time to a much greater extent compared to the oldest participants, suggesting both greater activation and impaired attenuation of immune signaling with advanced age.

### Age-dependent differences in cytokine and chemokine levels upon hospitalization and over time

The impact of aging on immune signaling in COVID-19 was also evident at the protein level. Analysis of proximity extension assay (Olink) protein data from serum samples identified 43 proteins that significantly correlated with age at the time of hospital admission (Fig. 5a, b, Supp. Fig. 7a). Of these, 31 increased with age, and the protein with the greatest effect size was CXCL9, a T-cell chemoattractant induced by IFN-γ and produced by neutrophils and macrophages^6^. Twelve proteins significantly decreased with age, including TNFSF11, which is involved in the regulation of T cell-dependent immune responses and group 2 innate lymphoid cell-mediated type 2 immunity^27^, and SIRT2, which may attenuate aging-associated inflammation through de-acetylation of the NLRP3 inflammasome^28^.

**Fig. 5:**
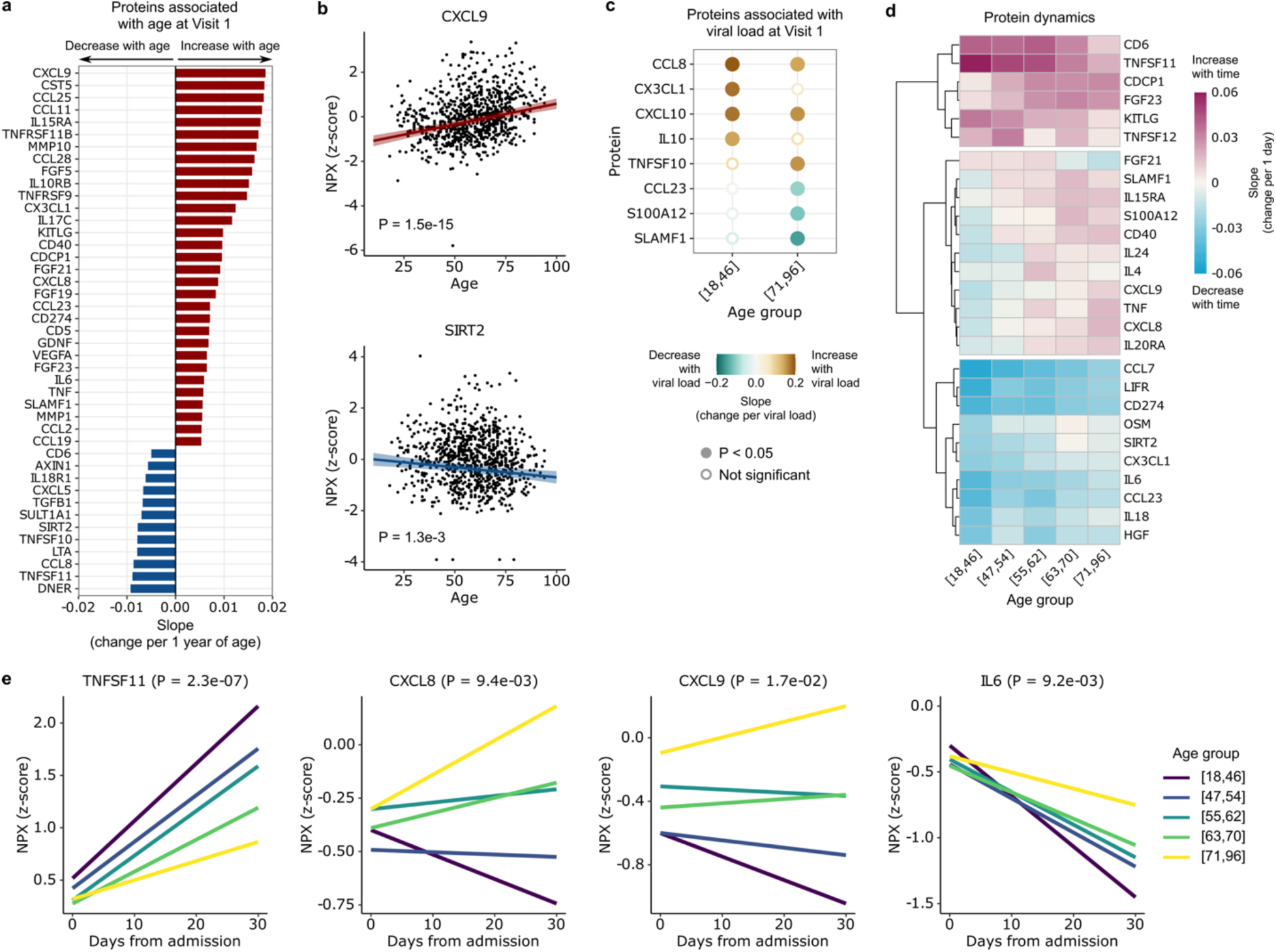
Aging leads to differences in cytokine and chemokine levels during COVID-19. (a) Bar plot highlighting proteins that are upregulated (red) or downregulated (blue) with age at Visit 1 (adjusted P < 0.05). (b) Scatter plots of the normalized protein expression (NPX) of representative proteins, CXCL9 and SIRT2, as a function of age. P values are calculated using linear regression and Benjamini-Hochberg correction. (c) Dot plot representing the slope of cytokine expression versus viral load in the youngest and oldest age quintiles, [18,46] and [71,96], respectively. (d) Heatmap depicting temporal slopes (i.e., change in protein expression per 1 day) of all cytokines that display age-dependent longitudinal dynamics (adjusted P < 0.05). (e) Plots showing the temporal dynamics of 4 example cytokines from (d). P values in (d, e) are calculated using linear mixed effects modeling and Benjamini-Hochberg correction.

Based on prior work^17^, we hypothesized that aging might affect the relationship between protein expression and viral load. Consistent with this idea, we identified eight cytokines and chemokines whose expression levels correlated with SARS-CoV-2 viral load (Fig. 5c), and observed differences in this relationship between the oldest and youngest age groups. For instance, expression of IL10, a key anti-inflammatory cytokine, increased more strongly in younger versus older adults in response to viral load. CX3CL1, a chemoattractant of T cells and monocytes, exhibited a similar relationship (Fig. 5c).

We next evaluated the longitudinal dynamics of cytokine/chemokine expression in the serum after hospitalization (Fig. 5d). The expression of several cytokines, such as TNFSF11, increased steeply over time in younger adults but lagged in the oldest adults (Fig. 5e, Supp. Fig. 7b). Conversely, the expression of several proinflammatory cytokines and chemokines such as CXCL8, CXCL9 and IL6 decreased rapidly over time in younger adults, while in the oldest adults expression increased over time (CXCL8, CXCL9) or declined more slowly (IL6) (Fig. 5e).

### Age-dependent changes in respiratory tract gene expression and the airway microbiome

We next asked whether aging was associated with changes in host gene expression and the upper airway microbiome (including virome) using nasal swab metatranscriptomics. We identified 913 host genes that were significantly associated with age (Fig. 6a), representing several key immune signaling pathways (Fig. 6b). TLR signaling, which plays an important role in microbial recognition, was upregulated with age, as were genes related to IFN-α/β, IL1, IL4, IL13, IL10, IL17, and caspase activation signaling. In contrast, T cell-related pathways (TCR signaling, co-stimulation by the CD28 family, and PD1 signaling) were downregulated with age, similar to our observations in peripheral blood. *In silico* prediction of upstream cytokine activation states demonstrated age-related activation of TNF, IL6, IFN-γ, IL1A/B, IL22 and CSF1 (Fig. 6c).

**Fig. 6:**
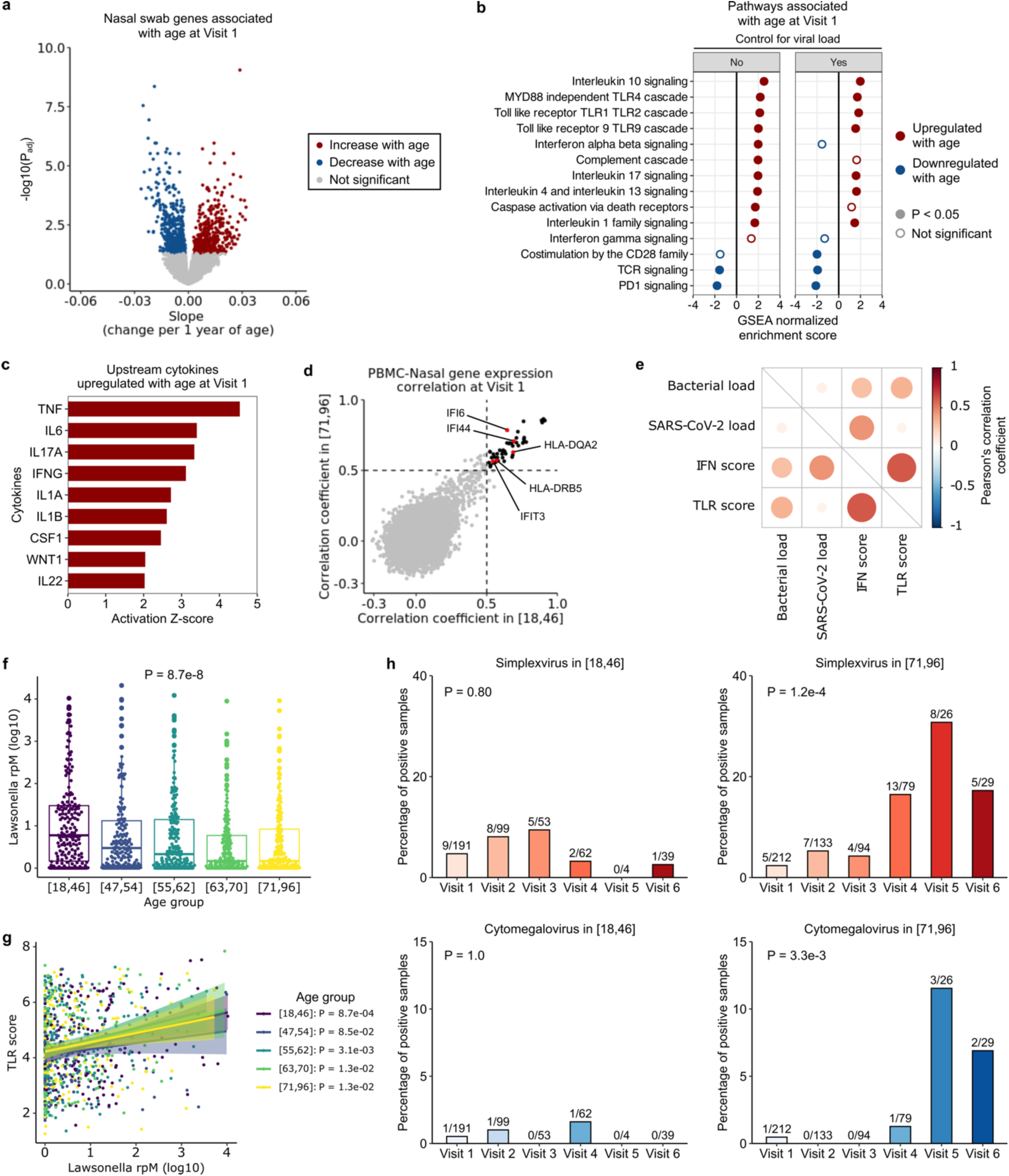
Aging changes upper respiratory tract gene expression and the airway microbiome in COVID-19. (a) Volcano plot depicting genes associated with age at Visit 1 in nasal swab metatranscriptomics data. (b) Normalized enrichment scores of select Reactome pathways associated with age at Visit 1, with (right) and without (left) controlling for viral load, in nasal samples. (c) Bar plot depicting cytokines predicted by Ingenuity Pathway Analysis to be upregulated with age in nasal samples. (d) Scatter plot depicting the Pearson’s correlation coefficient of gene expression between PBMC and nasal samples. Each dot indicates the correlation coefficient between PBMC expression and nasal expression of a gene, in the youngest (x-axis) and oldest (y-axis) age group. The black dots mark the genes with correlation coefficients > 0.5 in both age groups (n = 52 genes). (e) Dot plot highlighting correlations between SARS-CoV-2 viral load (log-transformed reads per million (rpM)), total bacterial abundance (log-transformed rpM), interferon stimulated gene (ISG) expression score and Toll like receptor (TLR) gene expression score. (f) Relative abundance of *Lawsonella* (rpM) across the age quintiles. In (f, g), P values were calculated with one-way ANOVA test. (g) Correlation between *Lawsonella* relative abundance and TLR gene expression across the age quintiles. P values were calculated using the test of association with Pearson’s correlation coefficient and adjusted with Benjamini-Hochberg correction. (h) Percentages of cases with herpes simplex virus (HSV) or cytomegalovirus (CMV) transcript detection in the youngest versus oldest age quintiles. The number on top of each bar indicates the number of positive cases over the number of total samples. P-values were calculated by Fisher exact test.

Our study design enabled assessment of inflammatory pathways across anatomic compartments. Thus, we were interested in the extent to which gene expression in the blood and the upper respiratory tract was coordinated. To this end, we calculated the Pearson’s correlation coefficients of gene expression between matched PBMC and nasal samples in the youngest group, and separately in the oldest group. We found 52 genes that had relatively high correlation coefficients in both groups, in particular those related to type I IFN signaling (e.g., *IFI6*, *IFI44, IFIT3*) and antigen presentation (HLA genes) (Fig. 6d).

As TLR pathways in the airway were strongly upregulated with age, we asked whether this could be due to differences in SARS-CoV-2 viral load and/or the nasal microbiome. We found that at Visit 1, the total bacterial load correlated with both ISG and TLR gene expression, while SARS-CoV-2 viral load only correlated with ISG expression (Fig. 6e). We thus considered whether variations in bacterial load across the age span might explain the observed age-related TLR signaling differences, however no variation was found (Supp. Fig. 8).

We also considered whether age-related differences in specific taxa within the airway microbiome might contribute to the aforementioned differences in TLR signaling. Metatranscriptomic analysis identified only one significant genus, *Lawsonella*, whose abundance decreased with age (Fig. 6f). *Lawsonella* abundance positively correlated with TLR gene expression across all age groups, however, demonstrating that it did not account for the age-related upregulation in TLR signaling (Fig. 6g). Lastly, we evaluated the upper respiratory tract virome, and observed reactivation of herpes simplex virus and cytomegalovirus over the course of hospitalization in the oldest age quintile, but not in younger participants (Fig. 6h). This suggested that older adults may have less capacity to maintain innate immune control of latent viral infections.

### Relationships between aging, immune response, and COVID-19 severity

Previous studies have established that severe COVID-19 involves a dysregulated host response characterized by inappropriate activation of inflammatory and immunoregulatory pathways^29–31^. We therefore sought to examine the intersection of aging, COVID-19 severity, and host immune responses by assessing PBMC gene expression differences at Visit 1 between participants with mild/moderate (baseline respiratory severity ordinal scale^2^ (OS) 3-4) and severe (baseline OS 5-6) COVID-19, within the youngest and oldest age groups (Fig. 7a).

**Fig. 7:**
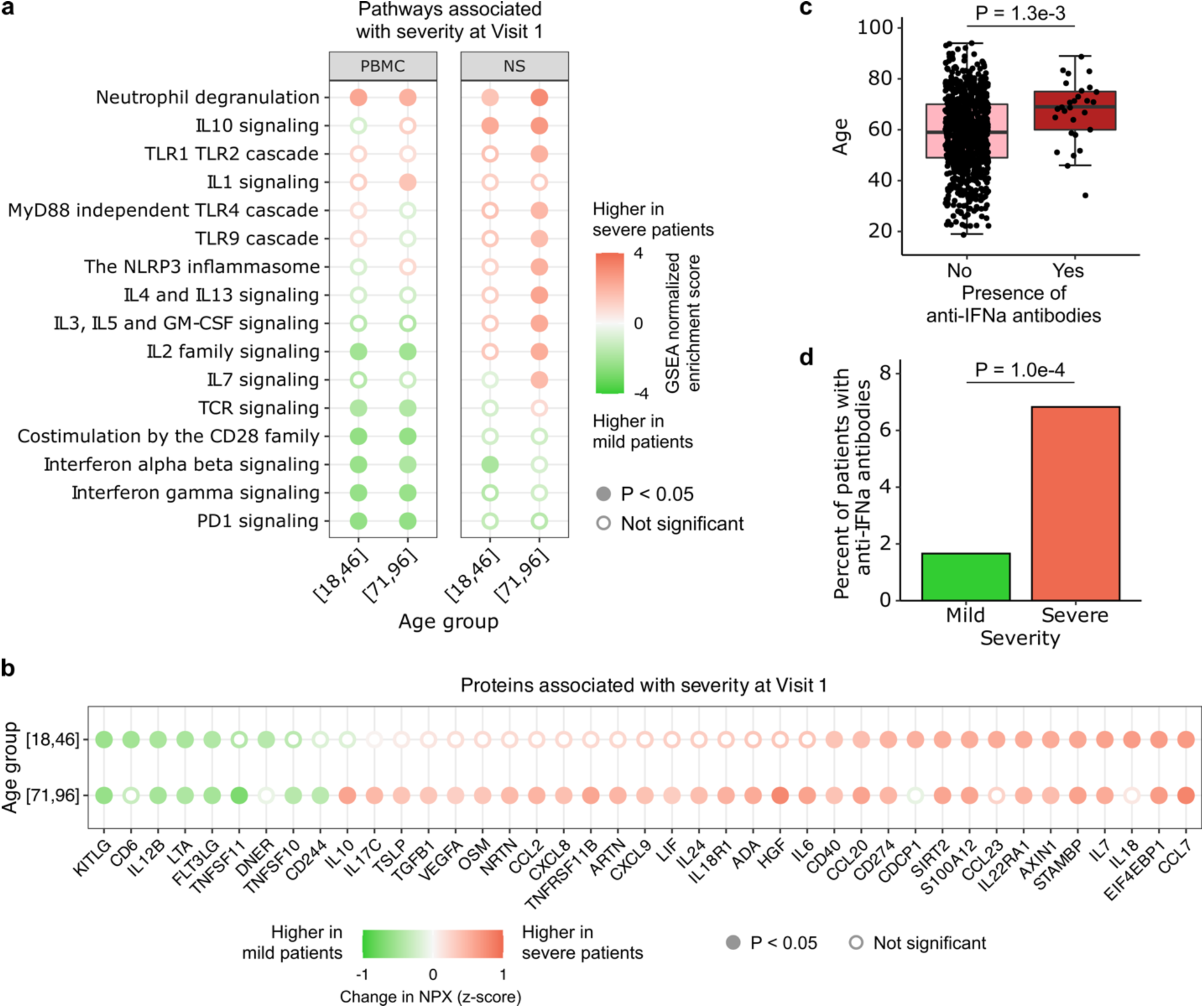
Aging and COVID-19 severity. (a, b) Dot plots highlighting a) select Reactome pathways in PBMC or nasal RNA-seq data, and b) serum proteins (Olink) that were upregulated in severe participants (baseline respiratory severity ordinal scale 5-6) compared to mild/moderate (ordinal scale 3-4) participants at Visit 1, stratified by age group (youngest or oldest). P values in (a, b) were calculated with linear modeling and Benjamini-Hochberg correction. (c) Box plot demonstrating association between age and presence of anti-IFN-α autoAbs in the 835 participants with available autoAb data at Visit 1. P value was calculated with the Wilcoxon rank-sum test. (d) Bar plot demonstrating the percentage of severe and mild/moderate participants who had anti-IFN-α antibodies (9/542 participants, 1.66% in mild/moderate; 20/293 participants, 6.83% in severe). P-value was calculated using the Chi-squared test.

Several immune signaling pathways were associated with disease severity in an age-dependent manner. For example, only in the oldest age quintile was severe COVID-19 associated with upregulation of IL3, IL5 and GM-CSF, IL4 and IL13, TLR, and NRLP3 inflammasome signaling pathways in the upper airway. Similarly in the blood, we found that the IL1 signaling pathway was only upregulated in severe COVID-19 in the oldest adults. We also identified several pathways that associated with COVID-19 severity independent of age. For example, in PBMCs from both the youngest and oldest participants, severe disease was associated with upregulation of neutrophil degranulation genes, and downregulation of pathways related to TCR, IFN-α/β, IFN-γ, IL2 and PD1 signaling.

Assessment at the protein level provided further insights regarding the immunological intersection of aging and COVID-19 severity (Fig. 7b). Notably, we found that the expression of several canonical proinflammatory cytokines and chemokines, such as IL6, oncostatin M (OSM), CXCL8 and CXCL9, was uniquely upregulated with disease severity in the oldest adults. Increased expression of the anti-inflammatory cytokines TGF-β1 and IL10 in severe disease was also specific to the oldest age quintile. Serum concentrations of several other proteins increased in severe disease independent of age, including CCL7, a leukocyte chemoattractant^32^, S100A12, a neutrophil-derived cytosolic pro-inflammatory protein^33^, and CD274 (PDL1), an immune checkpoint inhibitor. Similarly, we found that severity was associated with reduced expression of several cytokines regardless of age, including IL12B and LTA (TNF-β). We tested whether the differences in SARS-CoV-2 viral load could significantly influence the results, and found that they did not (Supp. Fig. 9).

Finally, we asked whether anti-IFN-α autoAb prevalence was associated with aging and COVID-19 severity. We found a significant positive correlation between age and anti-IFN-α autoAb prevalence at Visit 1 (Fig. 7c), and a greater prevalence of the autoAbs in participants with severe disease (Fig. 7d). The presence of anti-IFN-α autoAbs was also associated with impaired ISG expression (Supp. Fig. 10).

### Integrated analysis of protein and transcriptomics data

Finally, we sought to integrate findings across genes and proteins, and between the blood and airway compartments. Integrated network analysis of statistically significant age-associated proteins and age-associated genes from the blood identified three prominent nodes related to chemokine ligand (CCL) signaling, T cell signaling and the cell cycle (Fig. 8a, see Methods). Additionally, analysis of the ten most significant proteins and their immediately downstream genes illuminated the complex cross talk between several key immune mediators. For example, CXCL9 activates the genes *CXCR3* and *CXCR5,* and inhibits the gene *DPP4* (also known as *CD26*), which is upregulated on T cells after activation^34^ (Supp. Fig. 11). Of these, CXCL9 was positively associated with age, while the three downstream genes were negatively correlated with age.

**Fig. 8:**
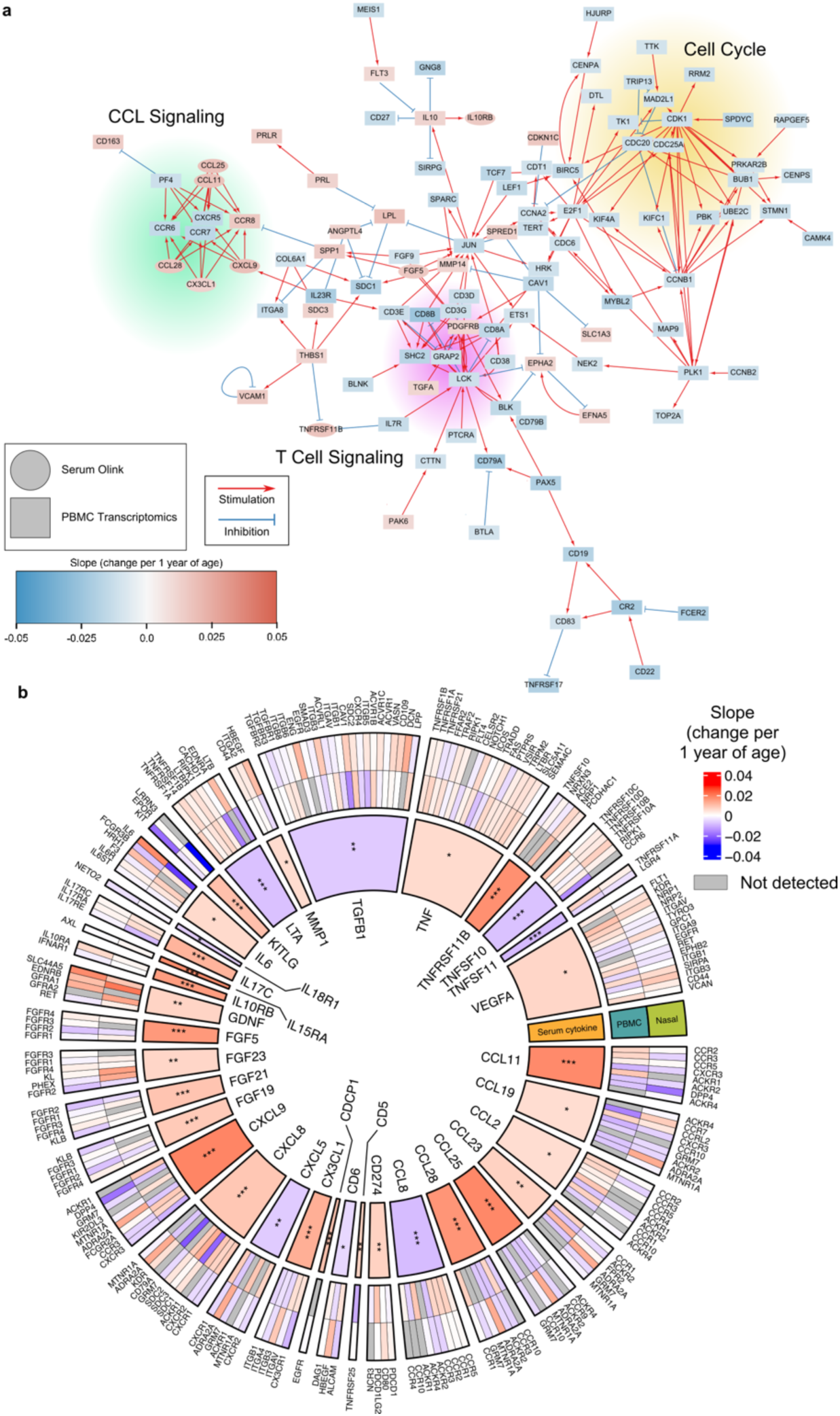
Integrated network analysis of serum cytokine/chemokine and PBMC and nasal transcriptomic data. (a) Network analysis of serum cytokines and PBMC genes significantly associated with age at Visit 1 using protein-protein interactions reported in Omnipath. (b) Analysis of ligand-receptor interactions from cytokine data, PBMC and nasal RNA-seq data. The inner most ring shows the significant cytokines from Visit 1 analysis and their magnitude of their average change per 1 year of age. The two outer rings illustrated genes that encode known receptors for each cytokine and their associated change per one year of age. P values for the cytokines were calculated using linear models and Benjamini-Hochberg correction. *P<0.05, **P<0.01, ***P<0.001.

To investigate how aging could potentially affect ligand-receptor interactions in both the blood and airway, we studied ligands in our protein data that were significantly associated with age at Visit 1, and examined the expression of the genes that encoded their cognate receptors. This analysis further highlighted transcriptomic/proteomic concordance and discordance (Fig. 8b). For example, aging was associated with increased expression of serum CXCL9 and CCL11, but decreased expression of *DPP4*, which encodes a receptor for these ligands. In contrast, the expression of both TNF ligand and its receptors (e.g., *TNFRSF1A*, *TNFRSF1B*, *LTBR*) were positively associated with age.

## Discussion

An effective host response to viral infection depends on potent early innate immune activation, engagement of adaptive immune effectors, and then, upon effective viral clearance, attenuation of this inflammatory signaling to prevent excessive tissue and pathologic consequences^35^. We observed age-dependent dysregulation of this program at the transcriptional, protein and cellular levels, manifesting in an imbalance of inflammatory responses over the course of hospitalization. Our results identify discrete innate and adaptive immune signaling pathways which are altered with age, suggesting potential targets for therapeutic intervention.

The role of type I IFN signaling in age-related immune dysregulation during COVID-19 has remained unclear, with some reports suggesting impaired induction of ISGs^7,15,24^ and others demonstrating the contrary^12,25^. We found that older age was associated with increased type I IFN signaling in both the blood and respiratory tract, but that the relationship was principally driven by differences in SARS-CoV-2 viral load. In contrast, IFN-γ signaling, which is associated with poor prognosis in COVID-19 participants^36^, was significantly upregulated with age independent of viral load.

Several factors likely contribute to higher SARS-CoV-2 viral loads in older adults, including impaired T and B cell immunity and impaired MHC antigen presentation, each of which we observed at the transcriptional, protein and cellular levels in our dataset. Delayed viral clearance due to these age-related factors could facilitate the evolution of novel SARS-CoV-2 variants^37,38^. Therefore, it is possible that coronavirus evolution could occur inside the host to a greater extent in older adults compared to the young, although our study was not designed to test this possibility.

Older adults had lower proportions of naïve CD8+ T cells and γδ T cells, which contribute to effective clearance of viral pathogens^24,39,40^. Terminally differentiated/exhausted NK cells, which are associated with severe COVID-19^41^, were more prevalent in older adults, as previously observed^12^, as were central memory CD8+ T cells. Impaired T cell signaling in older adults was also observed at the transcriptional level in both the blood and the upper respiratory tract upon hospitalization. Longitudinal analyses demonstrated attenuated expression dynamics of TCR signaling-related genes in the older participants’ blood samples (Fig. 3g, h). We also observed differences in the relationship between viral load and cytokines important for T cell recruitment in the oldest versus youngest adults, such as the chemokine CX3CL1.

Younger adults exhibited a much more robust induction of MHC II gene expression over the course of hospitalization. This is consistent with a previous study that reported *HLA-DR* expression increases over time following symptom onset in young COVID-19 participants, but not in older ones^12^. We also found that expression of MHC I genes decreased more rapidly post-hospitalization in the oldest versus the youngest adults. Given that SARS-CoV-2 can subvert immune responses by reducing MHC I surface levels in infected cells^42^, our results suggest that older participants may be even more vulnerable to this viral immune evasion mechanism.

Evidence of impaired B cell immunity was also observed in the older participants, consistent with prior studies^12,24^. Age was associated with reduced expression of genes involved in BCR signaling at the time of hospitalization. Furthermore, we observed lower proportions of naïve B cells and plasmablasts in the oldest adults. Functional ramifications of this were evident in decreased anti-SARS-CoV-2 RBD Ab levels, both upon hospitalization and when assessed longitudinally over 28 days.

Effective modulation of inflammatory responses is critical for restoration of immune homeostasis and mitigation of excessive tissue damage. We found consistent evidence of prolonged, potentially pathologic inflammatory responses in the oldest adults from transcriptomic and proteomic analyses. For instance, upon hospitalization, proinflammatory cytokines such as TNF, IL6, CXCL8, and CXCL9 were higher in the older participants, and continued to increase over time. In contrast, these cytokines decreased over time in the younger participants. Our results suggest that age-related changes may exacerbate the overexuberant inflammatory signaling in severe COVID-19, an early hypothesis^12,24^ that has not yet been validated in a large observational cohort.

The oldest adults in our cohort had evidence of HSV and CMV reactivation in the airway over the first 28 days after hospitalization. This may reflect impaired antiviral immune defenses in older adults exacerbated in the context of SARS-CoV-2 challenge. Furthermore, reactivation of latent Herpesviridae may contribute to excessive inflammatory responses observed in the older adults, as has been described in participants with human immunodeficiency virus (HIV) infection^43^.

Advanced age was also associated with upregulation of TLR signaling genes in both the airway and the blood, independent of SARS-CoV-2 viral load (Fig. 3e, 5b). We found that airway bacterial load correlated with TLR expression independent of age, and compositional differences in the microbiome across age groups did not explain this association, suggesting that age-related increases in TLR gene expression were caused by microbe-independent factors. Consistent with this idea are studies demonstrating that upregulation of innate immune receptors, including TLRs, could be an intrinsic feature of inflammaging^24,44^.

Severe COVID-19 is characterized by dysregulated, pathologic inflammatory responses^29,45,46^. We found that aging was associated with higher expression of several signaling pathways previously implicated in this pathologic immune dysregulation. For instance, in the oldest adults, severe COVID-19 was uniquely associated with impaired systemic Type 1 T helper cell (IL2, GM-CSF) and Type 2 T helper cell (IL5) responses, juxtaposed against hyperactivation of proinflammatory cytokines such as IL6, OSM, CXCL8, and CXCL9. In the airway, severe COVID-19 in the oldest age group led to greater NLRP3 inflammasome and TLR activation compared to the youngest group. These differences raise the possibility that older adults with severe COVID-19 may respond differently, and perhaps more favorably, to immunomodulatory therapies directed at certain inflammatory cytokines.

We also found that many features of severity-associated immune dysregulation were conserved across the lifespan, including an impairment in type I IFN signaling in severe disease. While presence of anti-IFN-α autoAbs was associated with impaired ISG expression and increased COVID-19 severity, they were detected in < 7% of adults in the oldest age quintile, suggesting a potentially important, but overall limited contribution to aging-associated COVID-19 severity relative to other immunological factors.

Our study is the largest molecular assessment of aging and COVID-19 to date (n = 1,031 participants at 20 hospitals across the United States), and one of the few to perform an integrated assessment of both immune and microbial features, allowing for identification of aging-related changes at a scale not previously achieved. We conducted multiomic, host/microbe systems immunoprofiling to assess the longitudinal dynamics of immune responses at the cellular, transcriptional and protein level in both the blood and airway. In addition, we add to the COVID-19 aging literature by integrating immunological analyses with assessment of both viral and microbiome dynamics over the course of hospitalization.

Our findings may have implications for age-specific COVID-19 therapeutic approaches. For example, a longer duration of antiviral therapy in older adults may be needed to achieve sufficient viral clearance for infection resolution compared to younger participants, and immunotherapy regimens may be particularly beneficial in older age, given impaired B cell responses. In addition, the optimal timing and use of immunomodulatory therapies (such as corticosteroids) may differ across the age spectrum, given the need to maximize control of inflammation without compromising the immune response to infection^47,48^.

Limitations of our study include the lack of a concurrently enrolled SARS-CoV-2-negative control group, and the lack of a non-hospitalized COVID-19 group. To partially address the first limitation, we analyzed publicly available gene expression datasets to incorporate findings from unrelated, healthy cohorts^5^. Participants in our current study were enrolled prior to the introduction of SARS-CoV-2 vaccines, and age-related differences in host immune responses may differ from a contemporary cohort due to variation in both vaccination status and the circulating SARS-CoV-2 variants. While this aspect limits extrapolation of our findings to immunized older adults, the naïve state of our study population was also a strength, as our results are not confounded by prior vaccination or infection, providing a window into age-related differences in immune response to a novel emerging viral respiratory pathogen.

In summary, we find that aging has marked impacts on host immune and viral dynamics in both recognized and novel ways in hospitalized participants with COVID-19. Notably, older adults exhibited impaired viral clearance, dysregulated immune signaling, and persistent and presumably pathologic activation of proinflammatory genes and cytokines.

## Materials and methods

### Patient enrollment and sample collection

This study leveraged data from the IMPACC cohort^2,23^, which enrolled participants from 20 hospitals across 15 medical centers in the United States between May 5th, 2020 and March 19th, 2021. Eligible participants were participants hospitalized with SARS-CoV-2 infection confirmed by RT-PCR and symptoms or signs consistent with COVID-19. The detailed study design and schedule for clinical data and biologic sample collection, and shared core platform assessments were previously described^23,30^. Detailed clinical assessments and sampling of blood and upper respiratory tract were performed within approximately 72 hours of hospitalization (Visit 1), and on approximately Days 4, 7, 14, 21, 28 after hospital admission. As previously described^23^, biological sample collection and processing followed a standard protocol utilized by every participating academic institution.

### Ethics

NIAID staff conferred with the Department of Health and Human Services Office for Human Research Protections (OHRP) regarding potential applicability of the public health surveillance exception [45CFR46.102(l)(2)] to the IMPACC study protocol. OHRP concurred that the study satisfied criteria for the public health surveillance exception, and the IMPACC study team sent the study protocol, and participant information sheet for review, and assessment to institutional review boards (IRBs) at participating institutions. Twelve institutions elected to conduct the study as public health surveillance, while three sites with prior IRB-approved biobanking protocols elected to integrate and conduct IMPACC under their institutional protocols (University of Texas at Austin, IRB 2020-04-0117; University of California San Francisco, IRB 20-30497; Case Western reserve university, IRB STUDY20200573) with informed consent requirements. Participants enrolled under the public health surveillance exclusion were provided information sheets describing the study, samples to be collected, and plans for data de-identification, and use. Those that requested not to participate after reviewing the information sheet were not enrolled. In addition, participants did not receive compensation for study participation while inpatient, and subsequently were offered compensation during outpatient follow-ups.

### Common statistical analyses framework

All raw data was obtained from the IMPACC study and are publicly available^2,23^. QC was performed by the IMPACC study as previously reported^2,23^. All data analyses were done in R v4.0.2. For each data type, we investigated the behavior of features both at Visit 1 (within 72 hours of hospital admission for most of the participants) and longitudinally for scheduled visits (Visits 1-6, up to 30 days post-hospital admission, both inpatient and outpatient samples, and excluding escalation samples). For Visit 1 analyses, we used linear modeling with age as a continuous variable and controlled for sex and baseline respiratory severity. Severity was assessed using a previously described 7-point severity ordinal scale (OS) based on degree of respiratory illness at the time of sampling^2^.

In the longitudinal analyses, we used age quintiles ([18,46], [47,54], [55,62], [63,70] and [71,96]), and controlled for sex and disease severity trajectory group (TG), a previously defined metric of COVID-19 severity over time. Clinical trajectory groups were previously identified and assigned to all participants in this study^2^. For longitudinal analysis of SARS-CoV-2 nasal viral load and serum anti-Spike IgG, we used generalized additive models with mixed effects from the package gamm4 (v0.2.6) to evaluate the effects of age while controlling for sex and TG. Generalized additive modeling was preferred for these features due to their non-linear trajectories as previously reported. For all other data types, we used linear mixed effects models from the package lme4 (v1.1.25). P values in all analyses were adjusted with Benjamini-Hochberg correction.

### Analysis of nasal metagenomics data

Taxonomic alignments from nasal metagenomics data were obtained from raw fastq files using the CZ-ID pipeline^49^, which first removes human sequences via subtractive alignment against human genome build 38, followed by quality and complexity filtering. Subsequently, reference-based taxonomic alignment at both the nucleotide and amino acid levels against sequences in the National Center for Biotechnology Information (NCBI) nucleotide (NT) and non-redundant (NR) databases, respectively, is carried out, followed by assembly of the reads matching each taxon. Taxa were aggregated to the genus level for analyses. For all analyses using SARS-CoV-2 viral load, log transformation of total reads per million (rpM) aligned to the Beta-coronavirus genus was used.

### Analysis of SARS-CoV-2 antibody titers

Antibody levels against the recombinant SARS-CoV-2 spike protein receptor-binding domain (RBD) were measured using a research-grade enzyme-linked immunosorbent assay (ELISA) as described^30^. The optical density (OD) was measured and the area under the curve was calculated, considering 0.15 OD as the cutoff.

Longitudinal analysis of SARS-CoV-2 viral load was performed using the gamm4 function from the gamm4 package (v0.2.6), using the following formula:

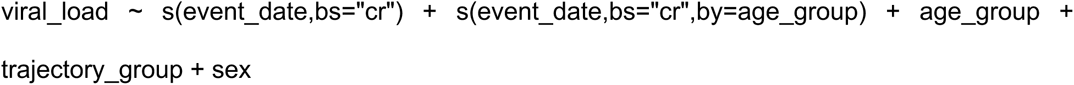

with random effects (1|site/pid). In the formula, viral_load is the log-transformed rpM counts of SARS-CoV-2 as measured by nasal metatranscriptomics, event_date was the number of days post hospitalization, age_group was the participant’s age quintile ([18,46], [47,54], [55,62], [63,70], [71,96]), TG was the participant’s trajectory group, site was the participant’s enrolment site and pid was the participant ID.

### Analysis of PBMC and nasal RNA-seq data

PBMC and nasal RNA-seq libraries were sequenced on a NovaSeq 6000 (Illumina) at 100 bp paired-end read length. The sequencing data was aligned using STAR aligner v2.4.2a and v.2.4.3^50^ and GRCh38 reference genome (Ensembl releases 91 and 100). Gene count tables were generated using htseq-count v0.4.1 and v0.4.2^51^.

For all RNA-seq analyses, we retained protein-coding genes that had a minimum of 10 counts in at least 20% of the samples. We normalized the gene counts using the voom function (normalize.method = “quantile”) from the limma package v3.46.0^52^, fitted a linear model for the gene expression with lmFit function (default settings), calculated the empirical Bayes statistics with ebayes function (default settings), and calculated the P values for differential expression with Benjamini-Hochberg multiple comparison correction. For Visit 1 analyses, we controlled for sex and severity OS at Visit 1, as well as log-transformed viral load in certain analyses when indicated. *In silico* prediction of upstream cytokine activation was performed with Qiagen’s Ingenuity Pathway Analysis software v01-21-03 (using default settings).

For the Visit 1 severity analysis, we defined mild/moderate participants as having baseline respiratory disease severity (OS) 3-4, and severe participants as having baseline OS 5-6, and limited to the youngest and oldest age quintiles. First, we normalized the gene counts with the voom function (normalize.method = “quantile”), and fitted a linear model for the gene expression using the lmFit function and the formula ∼ 0 + age_severity + sex, where age_severity is the combined categorical variable of participants’ ages (young, ≤ 46 years old, or old, ≥ 71 years old) and disease severity (mild or severe). With this parameterization, the age_severity variable has 4 levels: young_mild, young_severe, old_mild, and old_severe. To assess differences between severe and mild disease among young and old participants, we used the contrasts.fit function on the contrasts [young_severe – young_mild) and [old_severe – old_mild), respectively. Finally, we calculated the empirical Bayes statistics on the two contrasts with the ebayes function (default settings), and calculated the P values for differential expression with Benjamini-Hochberg correction.

For the longitudinal analysis, we restricted to hospitalized and outpatient samples that were collected up to 30 days post hospitalization, excluding samples collected during care escalation. We retained protein-coding genes with at least 10 counts in at least 20% of samples. Next, we normalized the gene counts using the voom function without adding any covariates, and modelled the normalized gene expression using linear mixed effects (LME) model with the lmer function from the lme4 package v1.1.25. Our LME model’s formula was:

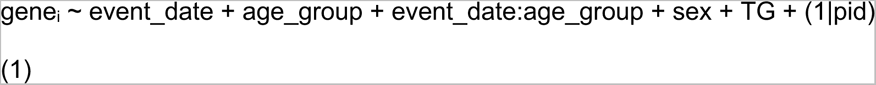

where gene_i_ was the normalized expression of gene i.

To calculate the P value of the interaction term between event_date and age_group, we used the anova function (test = “LRT”) to perform a likelihood ratio test between the model (1) above and the null model:

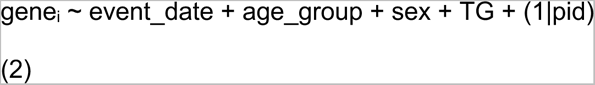

The P values from the likelihood ratio tests were then adjusted with Benjamini-Hochberg correction.

Significant genes from the longitudinal analysis of PBMC RNA-seq data were clustered with the pheatmap package v1.0.12, using the Euclidean distance metric and the Ward’s linkage (clustering_method = “ward.D2”). The TCR signaling genes and inflammatory genes were obtained from the corresponding Reactome and Hallmark pathways, respectively.

### Analysis of CyTOF data

Blood samples were quantified on the Fluidigm Helios mass cytometer, and the cell types were annotated using an automated annotation pipeline^30^. Prior to analysis, we removed cells identified as multiplets, debris, and those that were not identifiable with high confidence. Next, because neutrophils (CD16^hi^) were much more abundant than the other cell types (median 60% of all detected cells), they were also removed. Then, we normalized the cell type abundance for each participant by calculating the percentage of each cell type, adding a pseudocount of 1 to avoid taking the logarithm of zeros (the pseudocount is added even if the percentage is higher than zero), and computing its centered log ratio (CLR):

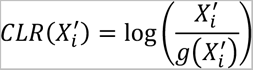

where log is the natural logarithm, and X_i_’ is the percentage of the cell type i:

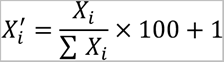

where X_i_ is the number of cells of cell type i in a participant.

For the Visit 1 analysis, we used a linear model to regress each cell type’s CLR-transformed abundance on age, while controlling for sex and OS. For the longitudinal analysis, we used a linear mixed effect model to model the CLR-transformed abundance. The formulae for the full and null models are identical to equations (1) and (2).

### Analysis of serum inflammatory protein (Olink) data

All samples were processed with the Olink multiplex assay inflammatory panels (Olink Proteomics), according to the manufacturer’s instructions and as previously described^30^. This inflammatory panel included 92 proteins associated with human inflammatory conditions. Target protein quantification was performed by real-time microfluidic qPCR via the Normalized Protein Expression (NPX) manager software. Data were normalized using internal controls in every sample, inter-plate control and negative controls, and correction factor and expressed as log2 scale proportional to the protein concentration. For additional quality control, we set any NPX measurements below the assay’s limit of detection (LOD) to zero. Next, we excluded proteins that were detected in fewer than 20% of samples, resulting in 84 proteins for analysis.

For the Visit 1 analysis, we standardized the NPX values and modeled them with linear regression on participants’ ages, controlling for sex and OS. For the severity analysis, we defined mild participants as baseline OS of 3-4, and severe participants as baseline OS of 5-6. We then set up two linear models, one for young participants (≤ 46 years old) and one for old participants (≥ 71 years old), to model the standardized NPX values on severity (mild or severe), while controlling for sex.

For the longitudinal analysis, we also standardized the NPX values, and used the LME models and the formulae in equations (1) and (2). Significant cytokines in the longitudinal analysis were clustered with the pheatmap package v1.0.12, using the Euclidean distance metric and the complete linkage. P values in all cytokine analyses were adjusted with Benjamini-Hochberg correction.

### Analysis of anti-IFN-α antibody presence and correlation with interferon-related gene expression

Samples were screened for anti-type I IFN autoAbs in a multiplex, particle-based assay as previously described^30^. Participant samples with a fluorescence intensity > 3 standard deviations above a mean of 1099 healthy controls at the earliest timepoint received were considered positive for anti-IFN Abs (> 1310 FI for IFN-α). To assess whether presence of anti-IFN-α Abs correlated with changes in IFN-related gene expression, we assessed expression of genes present in the REACTOME pathway “interferon alpha beta signaling” obtained from gsea-msigdb.org. All genes were assessed by linear regression using the formula lm(exp ∼ anti_IFNa + age + sex + viral_load), where exp was the normalized gene expression from PBMC data, antiIFNa was a binary variable indicating presence or absence of anti-IFN-α Abs, and viral_load was the log-transformed viral load measured by SARS-CoV-2 rpM from the nasal metagenomics data. P values were adjusted with Benjamini-Hochberg correction.

### Integrated analysis of serum cytokine, PBMC RNA-seq and nasal RNA-seq data

To integrate the serum cytokines/chemokines with the PBMC and nasal transcriptomic data, ligand/receptor pairs were retrieved from Omnipath, a database of known protein-protein interactions, using the R package OmnipathR to identify receptors and protein associates for ligands in the serum OLINK. The change per year of age was graphed for significant age-associated ligands in the serum OLINK and their respective receptors/interactive proteins for both transcriptomics using the R package ComplexHeatmap.

## Data Availability

All data are available under restricted access to comply with NIH Public data sharing policy for IRB-exempted public surveillance studies. Access can be obtained via AccessClinicalData@NIAID (https://accessclinicaldata.niaid.nih.gov/study-viewer/clinical_trials). More information can also be found under https://docs.immport.org/home/impaccslides/. Data files were previously generated (2, 36, 63) and are available at ImmPort under accession number SDY1760 and dbGAP accession number phs002686.v1.p1 (clinical data version 2021-11-11-frozen). The healthy control aging transcriptomic dataset was obtained from a supplementary data file from Peters et al.(5). All analysis code has been deposited at https://bitbucket.org/kleinstein/impacc-public-code/src/master/aging_manuscript/.

## List of Supplementary Materials

Supplementary Table 1. Clinical and demographic characteristics of the cohort at baseline (Visit 1).

Supplementary Figure 1. Comparison of viral load as measured by nasal swab qPCR and nasal swab RNA-seq (metatranscriptomics).

Supplementary Figure 2. Time since symptom onset across age groups at Visit 1. Supplementary Figure 3. Visit 1 analysis and longitudinal analysis of IgG levels. Supplementary Figure 4. Visit 1 analysis of CyTOF data while controlling for viral load.

Supplementary Figure 5. Comparison of PBMC RNA-seq data from this study to healthy control datasets, with differential gene expression analyses performed using age as a continuous variable.

Supplementary Figure 6. Plots of the dynamics of 6 example genes in PBMC samples.

Supplementary Figure 7. Effect of SARS-CoV-2 viral load on age-cytokine relationship at Visit 1, and the dynamics of 4 example cytokines.

Supplementary Figure 8. Bacterial load (reads per million, rpM) versus age quintiles.

Supplementary Figure 9. Aging and COVID-19 severity, analyses controlled for viral load.

Supplementary Figure 10. Expression of interferon-related genes in patients with or without anti-IFN-α antibodies at Visit 1.

Supplementary Figure 11. Network analysis of the top 10 significant serum proteins and their receptors and downstream signaling.

Supplementary Table 2. Prevalence of viral cases by age quintile over time in the nasal virome.

## CONSORTIA

**^#^The IMPACC Network**

**National Institute of Allergy and Infectious Diseases, National Institute of Health, Bethesda, MD 20814, USA:** Patrice M. Becker, Alison D. Augustine, Steven M. Holland, Lindsey B. Rosen, Serena Lee, Tatyana Vaysman

**Clinical and Data Coordinating Center (CDCC) Precision Vaccines Program, Boston Children’s Hospital, Boston, MA 02115, USA:** Al Ozonoff, Joann Diray-Arce, Jing Chen, Alvin Kho, Carly E. Milliren, Annmarie Hoch, Ana C. Chang, Kerry McEnaney, Brenda Barton, Claudia Lentucci, Maimouna Murphy, Mehmet Saluvan, Tanzia Shaheen, Shanshan Liu, Caitlin Syphurs, Marisa Albert, Arash Nemati Hayati, Robert Bryant, James Abraham

**Benaroya Research Institute, University of Washington, Seattle, WA 98101, USA: Matthew C. Altman, Naresh Doni Jayavelu, Scott Presnell, Bernard Kohr, Azlann Arnett**

**La Jolla Institute for Immunology, La Jolla, CA 92037, USA:** Bjoern Peters, Randi Vita, Kerstin Westendorf

**Knocean Inc. Toronto, ON M6P 2T3, Canada:** James A. Overton

**Precision Vaccines Program, Boston Children’s Hospital, Harvard Medical School, Boston, MA 02115, USA:** Ofer Levy, Hanno Steen, Patrick van Zalm, Benoit Fatou, Kinga Smolen, Arthur Viode, Simon van Haren, Meenakshi Jha

**Brigham and Women’s Hospital, Harvard Medical School, Boston, MA 02115, USA:** Lindsey. R. Baden, Kevin Mendez, Jessica Lasky-Su, Alexandra Tong, Rebecca Rooks

**Metabolon Inc, Morrisville, NC 27560, USA:** Scott R. Hutton, Greg Michelotti, Kari Wong

**Case Western Reserve University and University Hospitals of Cleveland, Cleveland, OH 44106, USA:** Rafick-Pierre Sekaly, Slim Fourati, Grace A. McComsey, Paul Harris, Scott Sieg, Susan Pereira Ribeiro

**Drexel University, Tower Health Hospital, Philadelphia, PA 19104, USA:** Charles B. Cairns, Elias K. Haddad, Michele A. Kutzler, Mariana Bernui, Gina Cusimano, Jennifer Connors, Kyra Woloszczuk, David Joyner, Carolyn Edwards, Edward Lin, Nataliya Melnyk, Debra L. Powell, James N. Kim, I. Michael Goonewardene, Brent Simmons, Cecilia M. Smith, Mark Martens, Brett Croen, Nicholas C. Semenza, Mathew R. Bell, Sara Furukawa, Renee McLin, George P Tegos, Brandon Rogowski, Nathan Mege, Kristen Ulring

**MyOwnMed Inc., Bethesda, MD 20817, USA:** Vicki Seyfert-Margolis

**Emory School of Medicine, Atlanta, GA 30322, USA:** Nadine Rouphael, Steven E. Bosinger, Arun K. Boddapati, Greg K. Tharp, Kathryn L. Pellegrini, Brandi Johnson, Bernadine Panganiban, Christopher Huerta, Evan J. Anderson, Hady Samaha, Jonathan Sevransky, Laurel Bristow, Elizabeth Beagle, David Cowan, Sydney Hamilton, Thomas Hodder

**Icahn School of Medicine at Mount Sinai, New York, NY 10029, USA:** Ana Fernandez-Sesma, Viviana Simon, Florian Krammer, Harm Van Bakel, Seunghee Kim-schulze, Ana Silvia Gonzalez-Reiche, Jingjing Qi, Brian Lee, Juan Manuel Carreño, Gagandeep Singh, Ariel Raskin, Johnstone Tcheou, Zain Khalil, Adriana van de Guchte, Keith Farrugia, Zenab Khan, Geoffrey Kelly, Komal Srivastava, Lily Eaker, Maria Carolina Bermúdez González, Lubbertus C.F. Mulder, Katherine Beach

**Immunai Inc. New York, NY 10016, USA:** Adeeb Rahman

**Oregon Health Sciences University, Portland, OR 97239, USA:** William B. Messer, Catherine L. Hough, Sarah Siegel, Peter Sullivan, Zhengchun Lu

**Stanford University School of Medicine, Palo Alto, CA 94305, USA:** Holden Maecker, Bali Pulendran, R. Kari C. Nadeau, Yael Rosenberg-Hasson, Michael Leipold, Natalia Sigal, Angela Rogers, Andrea Fernandez, Monali Manohar, Evan Do, Iris Chang

**David Geffen School of Medicine at the University of California Los Angeles, Los Angeles CA 90095, USA:** Elaine F. Reed, Joanna Schaenman, Ramin Salehi-Rad, Adreanne M. Rivera, Harry C. Pickering, Subha Sen, David Elashoff, Dawn C. Ward

**University of California San Francisco, San Francisco, CA 94115, USA:** David J. Erle, Carolyn S. Calfee, Carolyn M. Hendrickson, Kirsten N. Kangelaris, Viet Nguyen, Deanna Lee, Suzanna Chak, Rajani Ghale, Ana Gonzalez, Alejandra Jauregui, Carolyn Leroux, Luz Torres Altamirano, Ahmad Sadeed Rashid, Andrew Willmore, Prescott G. Woodruff, Matthew F. Krummel, Sidney Carrillo, Alyssa Ward, Charles R. Langelier, Ravi Patel, Michael Wilson, Ravi Dandekar, Bonny Alvarenga, Jayant Rajan, Walter Eckalbar, Andrew W. Schroeder, Gabriela K. Fragiadakis, Alexandra Tsitsiklis, Eran Mick, Yanedth Sanchez Guerrero, Rajani Ghale, Christina Love, Lenka Maliskova, Michael Adkisson

**Yale School of Medicine, New Haven, CT 06510, USA:** David A. Hafler, Ruth R. Montgomery, Albert C. Shaw, Steven H. Kleinstein, Jeremy Gygi, Shrikant Pawar, Anna Konstorum, Ernie Chen, Chris Cotsapas, Xiaomei Wang, Leqi Xu, Charles Dela Cruz, Akiko Iwasaki, Subhasis Mohanty, Allison Nelson, Yujiao Zhao, Shelli Farhadian, Hiromitsu Asashima

**Yale School of Public Health, New Haven, CT 06510, USA:** Denise Esserman, Leying Guan, Anderson Brito, Jessica Rothman, Nathan Grubaugh, Albert I. Ko

**Baylor College of Medicine and the Center for Translational Research on Inflammatory Diseases, Houston, TX 77030, USA:** David B. Corry, Farrah Kheradmand, Li-Zhen Song, Ebony Nelson

**Oklahoma University Health Sciences Center, Oklahoma City, OK 73104, USA:** Jordan P. Metcalf, Nelson I Agudelo Higuita, Lauren Sinko, J. Leland Booth

**University of Arizona, Tucson AZ 85721, USA:** Monica Kraft, Chris Bime, Jarrod Mosier, Heidi Erickson, Ron Schunk, Hiroki Kimura, Michelle Conway

**University of Florida, Gainesville, FL 32611, USA:** Mark A. Atkinson, Scott C. Brakenridge, Ricardo F. Ungaro, Brittany Roth Manning,

**University of Florida, Jacksonville, FL 32218, USA:** Jordan Oberhaus, Faheem W. Guirgis,

**University of South Florida, Tampa FL 33620, USA:** Brittney Borresen, Matthew L. Anderson

**University of Texas, Austin, TX 78712, USA:** Lauren I. R. Ehrlich, Esther Melamed, Cole Maguire, Justin F. Rousseau, Kerin C. Hurley, Janelle N. Geltman, Nadia Siles, Jacob E. Rogers

## ACKNOWLEDGMENTS

**Clinical and Data Coordinating Center:** Sanya Thomas, Mitchell Cooney, Shun Rao, Sofia Vignolo, Elena Morrocchi. **David Geffen School of Medicine at the University of California-Los Angeles:** Arash Naeim, Marianne Bernardo, Sarahmay Sanchez, Shannon Intluxay, Clara Magyar, Jenny Brook, Estefania Ramires-Sanchez, Megan Llamas, Claudia Perdomo, Clara E. Magyar, Jennifer A. Fulcher, and the UCLA Center for Pathology Research Services and the Pathology Research Portal. **Yale School of Medicine:** M. Catherine Muenker, Dimitri Duvilaire, Maxine Kuang, William Ruff, Khadir Raddassi, Denise Shepherd, Haowei Wang, Omkar Chaudhary, Syim Salahuddin, John Fournier, Michael Rainone, Maxine Kuang.

## Funding

The study was funded by the United States National Institutes of Health through the following grants: 5R01AI135803-03, 5U19AI118608-04, 5U19 AI128910-04, 5U19 AI089992, 4U19AI090023-11, 4U19AI118610-06, R01AI145835-01A1S1, 5U19AI062629-17, 5U19AI057229-17, 5U19AI125357-05, 5U19AI128913-03, 3U19AI077439-13, 5U54AI142766-03, 5R01AI104870-07, 3U19AI089992-09, 3U19AI128913-03, NIH-NIAID 3U19AI1289130 and U19AI128913-04S1 for EFR, R01 AI122220 for CC.

## Author contributions

C.R.L. conceived the idea for the project. H.V.P., A.T., C.P.M. and B.L. analyzed the data. E.K.H., P.M.B., S.K.S., J.C., A.H., H.P., P.v.Z., M.C.A., A.D.A., C.S.C., S.B., C.C., W.E., L.G., N.D.J., S.H.K., F.K., H.T.M., A.O., B.P., N.R., R.R.M., E.R., J.S., H.S., O.L. and J.D.A. provided guidance. H.V.P., A.T. and C.R.L. wrote the manuscript. All authors reviewed and edited the manuscript.

## Data and code availability

Data files are available at ImmPort under accession number SDY1760 and dbGAP accession number phs002686.v1.p1. All analysis code has been deposited at https://bitbucket.org/kleinstein/impacc-public-code/src/master/aging_manuscript/.

## Supplementary Materials

**Supplementary Figure 1.**
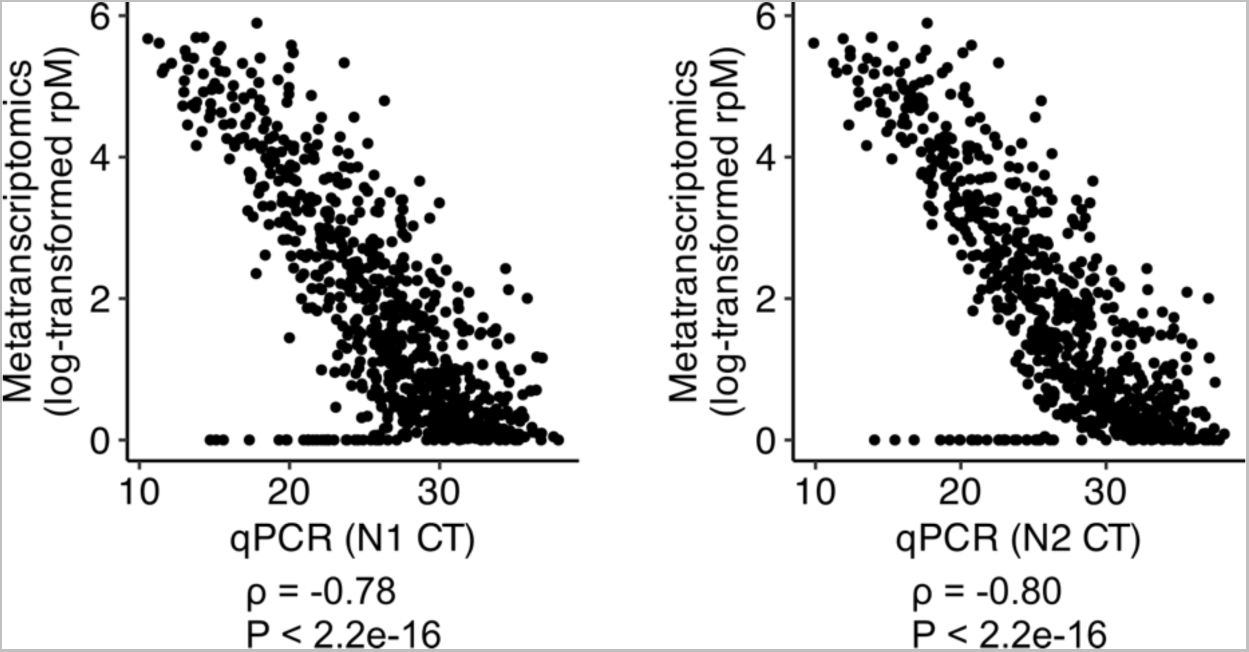
Comparison of viral load as measured by nasal swab qPCR and nasal swab RNA-seq (metatranscriptomics). The Pearson’s correlation coefficient and its P value are shown below each panel.

**Supplementary Figure 2.**
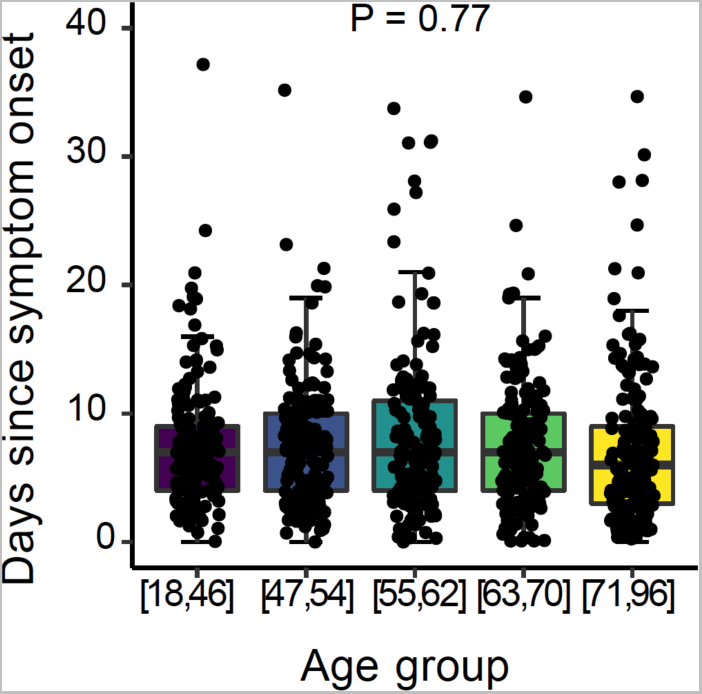
Time since symptom onset across age groups at Visit 1. Data is available for a subset of patients (n=796 of 963 Visit 1 samples). Two outliers with >40 days since symptom onset are excluded from the plot. P value is calculated by one-way ANOVA test.

**Supplementary Figure 3.**
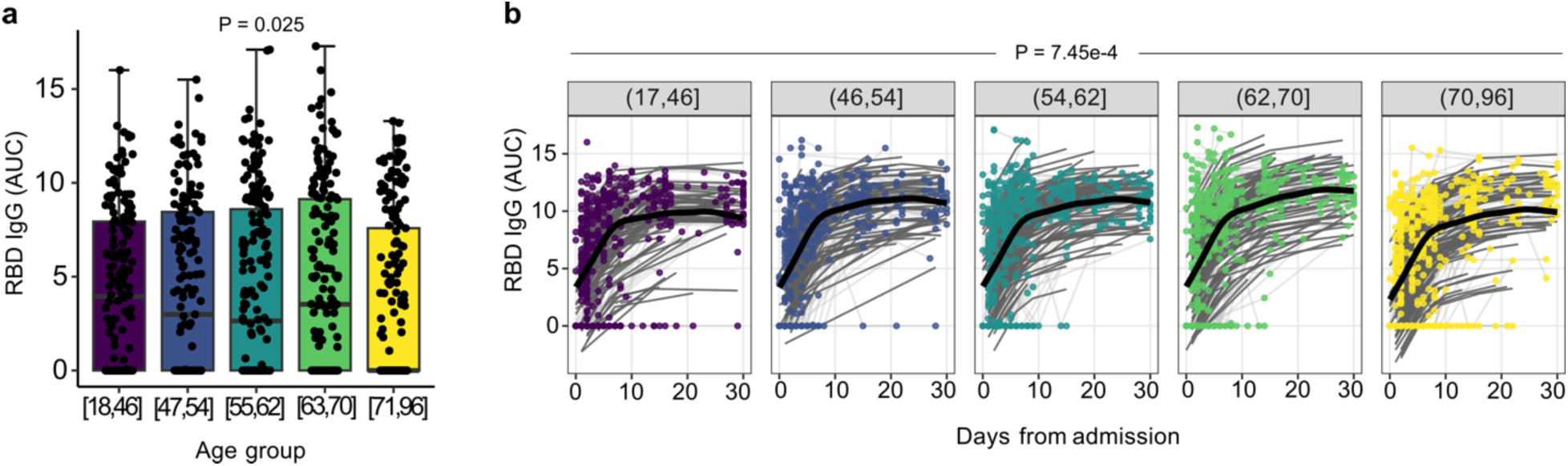
Visit 1 analysis and longitudinal analysis of IgG levels. (a) RBD IgG at visit 1 in each age group. P-value determined by likelihood ratio test. (b) RBD IgG levels, as measured by area under the curve (AUC, see methods), over time in each age group. P-value determined by a generalized additive mixed model. Values plotted represent the area under the curve of the optical density (OD).

**Supplementary Figure 4.**
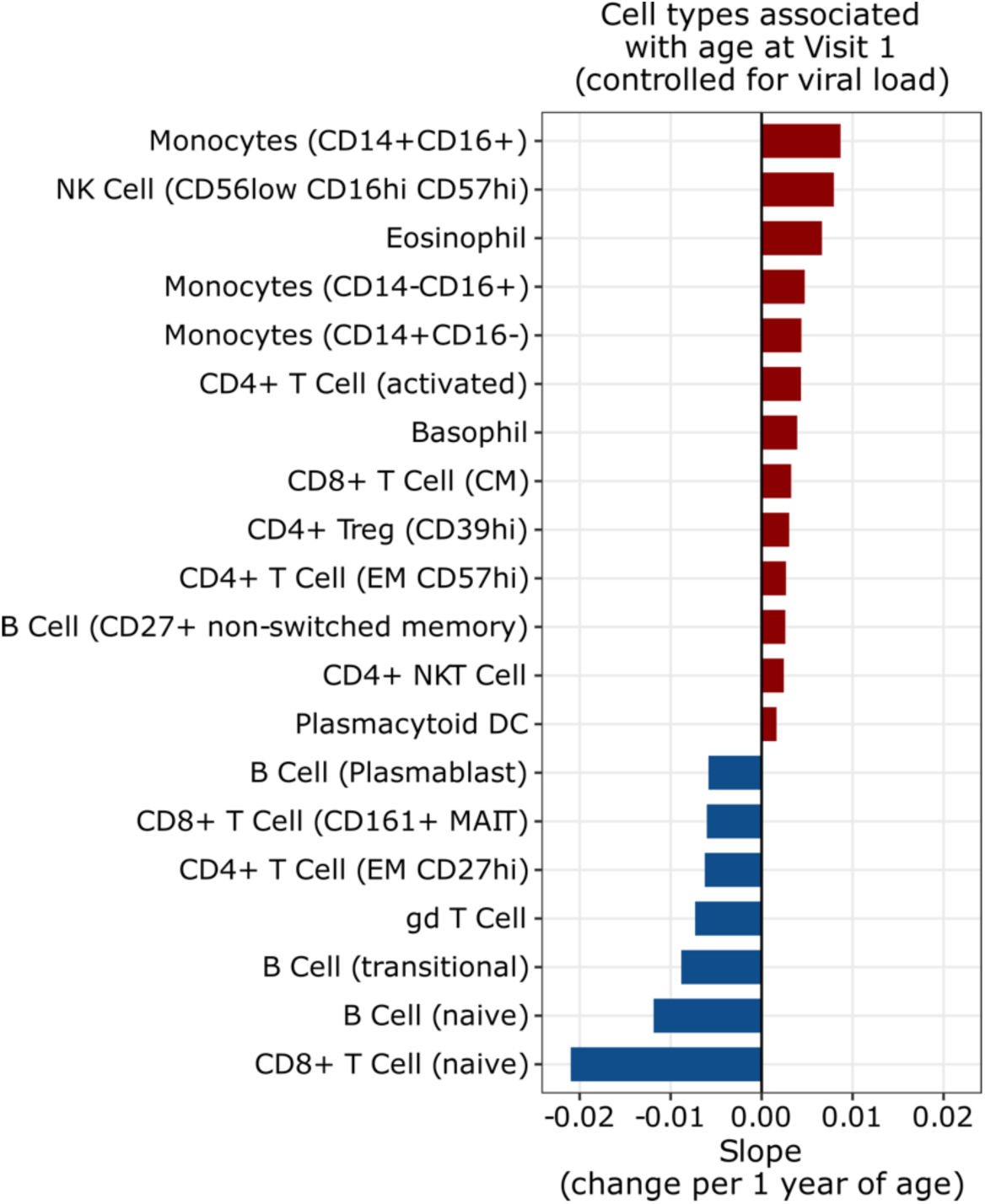
Visit 1 analysis of CyTOF data while controlling for viral load. Bar plot highlighting cell types that are significantly associated with age (P < 0.05, calculated with linear regression and Benjamini-Hochberg correction). Analogous to the analyses in Figure 3b, but controlled for viral load.

**Supplementary Figure 5.**
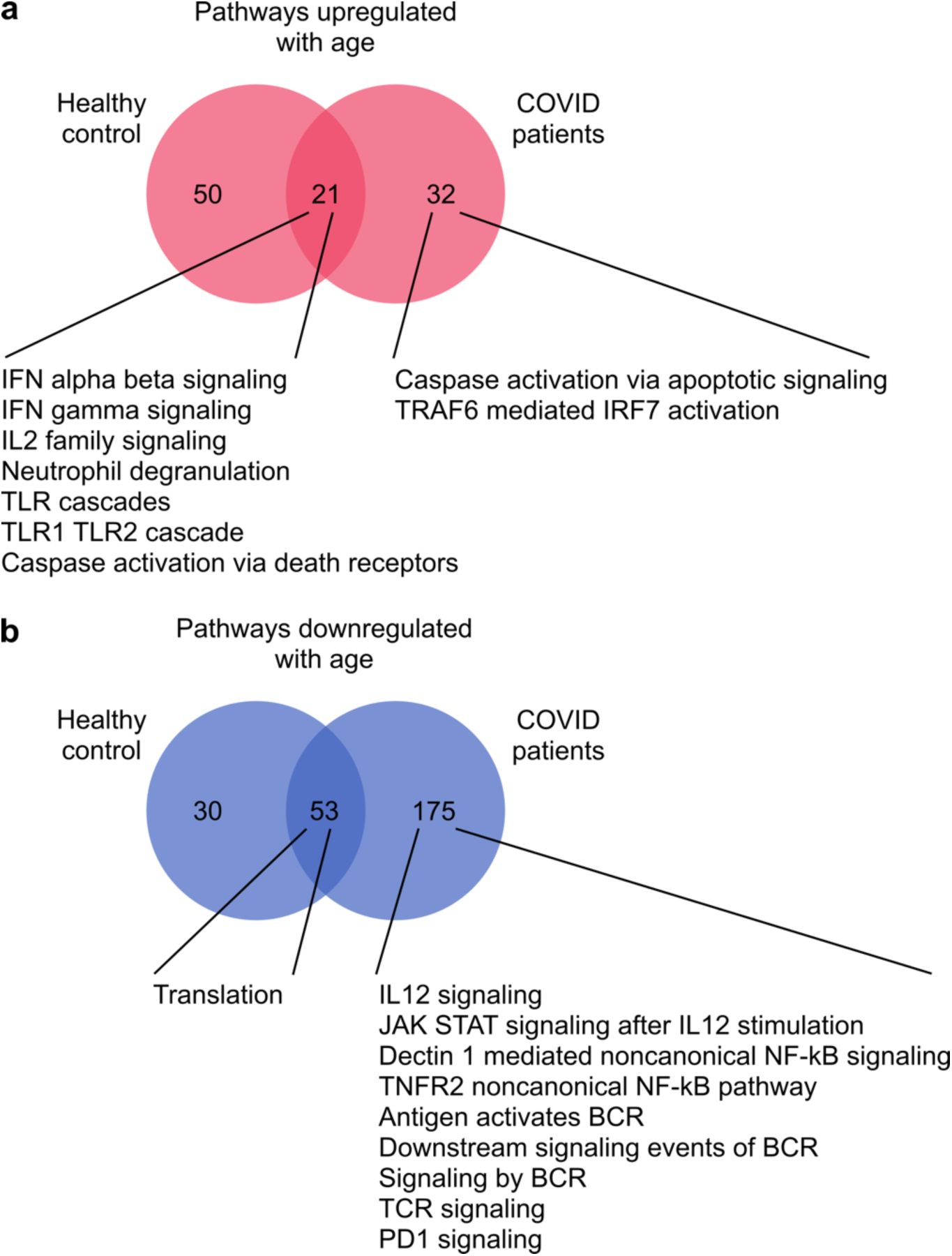
Comparison of PBMC RNA-seq data from this study to healthy control datasets^5^, with differential gene expression analyses performed using age as a continuous variable. (a, b) Venn diagrams of the Reactome pathways that are (a) upregulated and (b) downregulated with age. The numbers in the left circles indicate the number of pathways that are up- or down-regulated with age in the healthy control data only. The numbers in the right circles indicate the number of pathways that are up- or down-regulated with age in COVID-19 patients (our data) only. The numbers in the overlapping regions indicate the number of pathways that are up- or down-regulated with age in both healthy control and COVID-19 patients. Some examples of overlapping pathways, and of pathways that are associated with age in COVID-19 patients only are included under each Venn diagram.

**Supplementary Figure 6.**
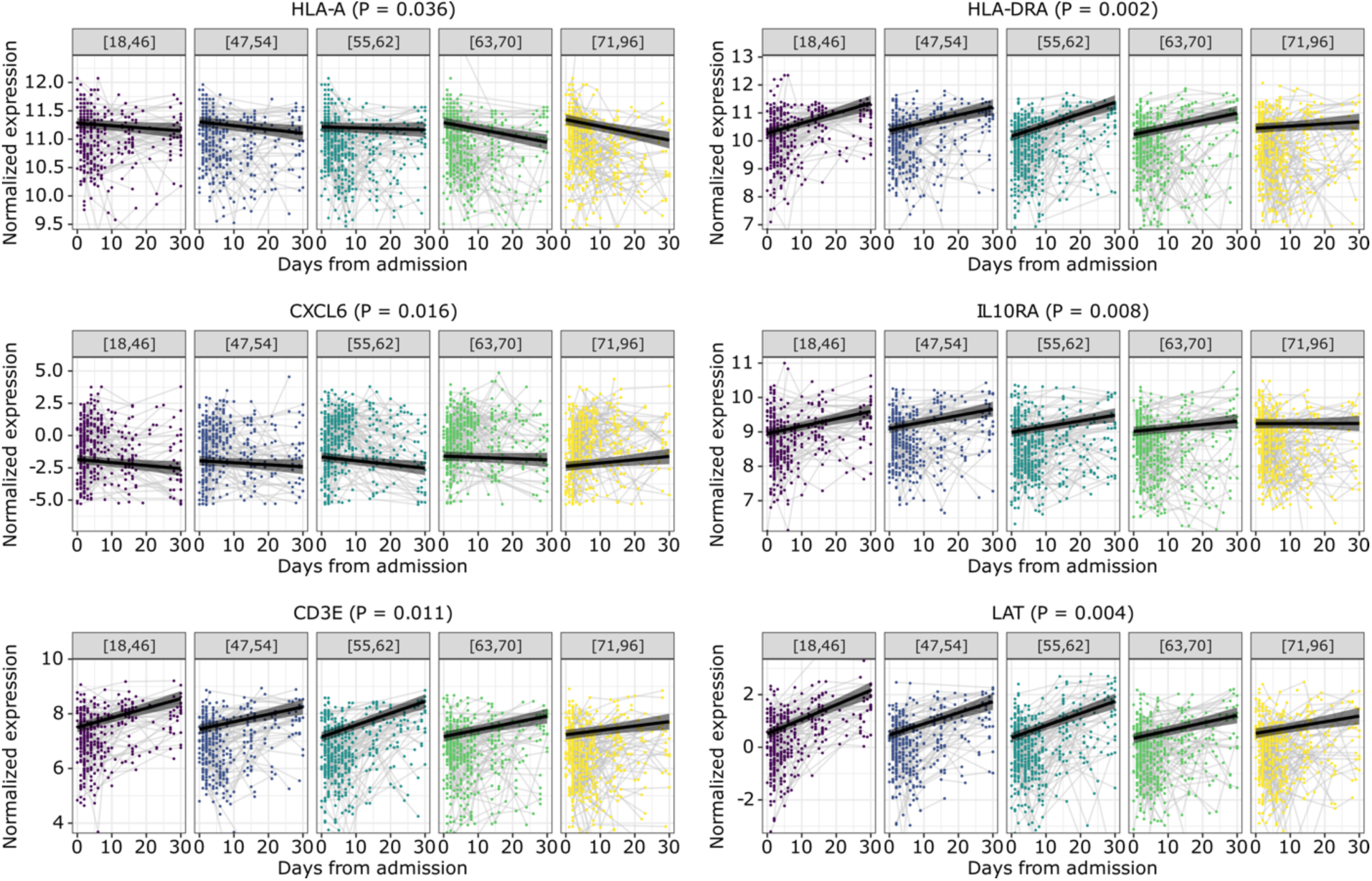
Plots of the dynamics of 6 example genes in PBMC samples. Black lines indicate the regression lines for the fixed effects of the linear mixed-effects model. The grey ribbons indicate the 95% confidence intervals of the regression lines. The y-axes were truncated at 1.5× the interquartile range below the first quartile and above the third quartile. P values are calculated using the likelihood ratio test and Benjamini-Hochberg correction.

**Supplementary Figure 7.**
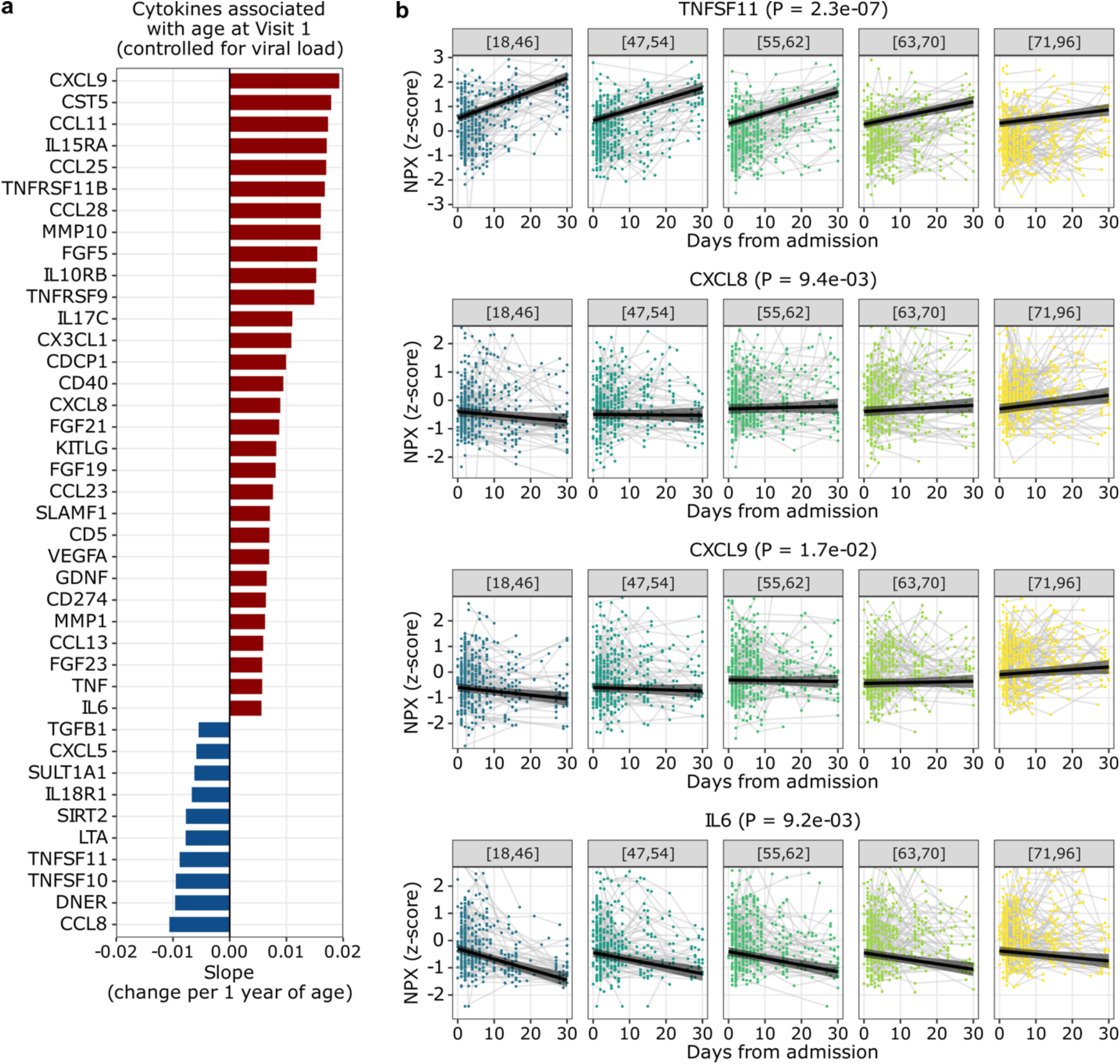
Effect of SARS-CoV-2 viral load on age-cytokine relationship at Visit 1, and the dynamics of 4 example cytokines. (a) Bar plot depicting cytokines associated with age (P < 0.05, calculated with linear regression and Benjamini-Hochberg correction), while controlling for SARS-CoV-2 viral load. Supplementary Figure 6a differs from Figure 4a in that the former controlled for viral load, while the latter did not. (b) Plots demonstrating the dynamics of four cytokines TNFSF11, CXCL8, CXCL9, and IL6. Black lines indicate the regression lines for the fixed effects of the linear mixed-effects model. The grey ribbons indicate the 95% confidence intervals of the regression lines. The y-axes were truncated at 1.5× the interquartile range below the first quartile and above the third quartile. P values were calculated using the likelihood ratio test and Benjamini-Hochberg correction.

**Supplementary Figure 8.**
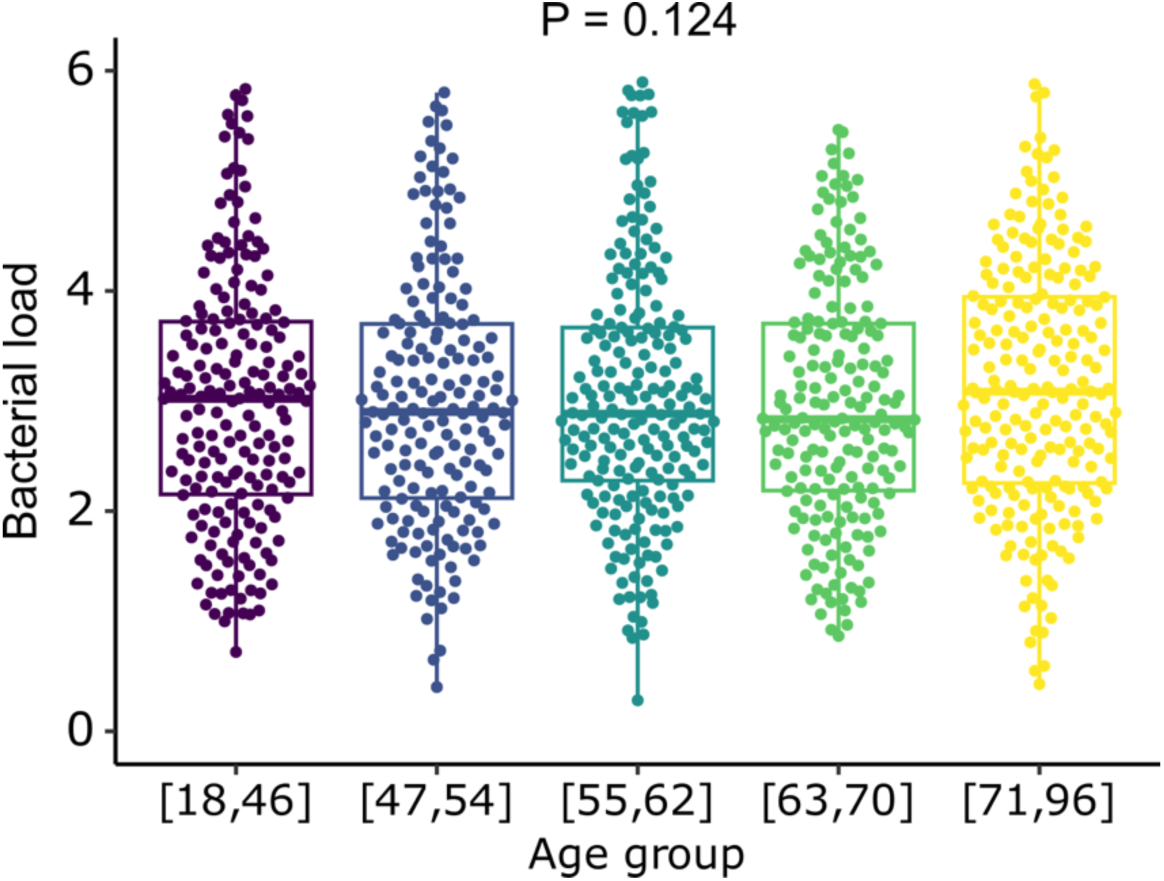
Bacterial load (reads per million, rpM) versus age quintiles. Total bacterial abundance (log-transformed rpM, as measured by nasal metatranscriptomics) in each of the age quintiles. P value was calculated with one-way ANOVA test.

**Supplementary Figure 9.**
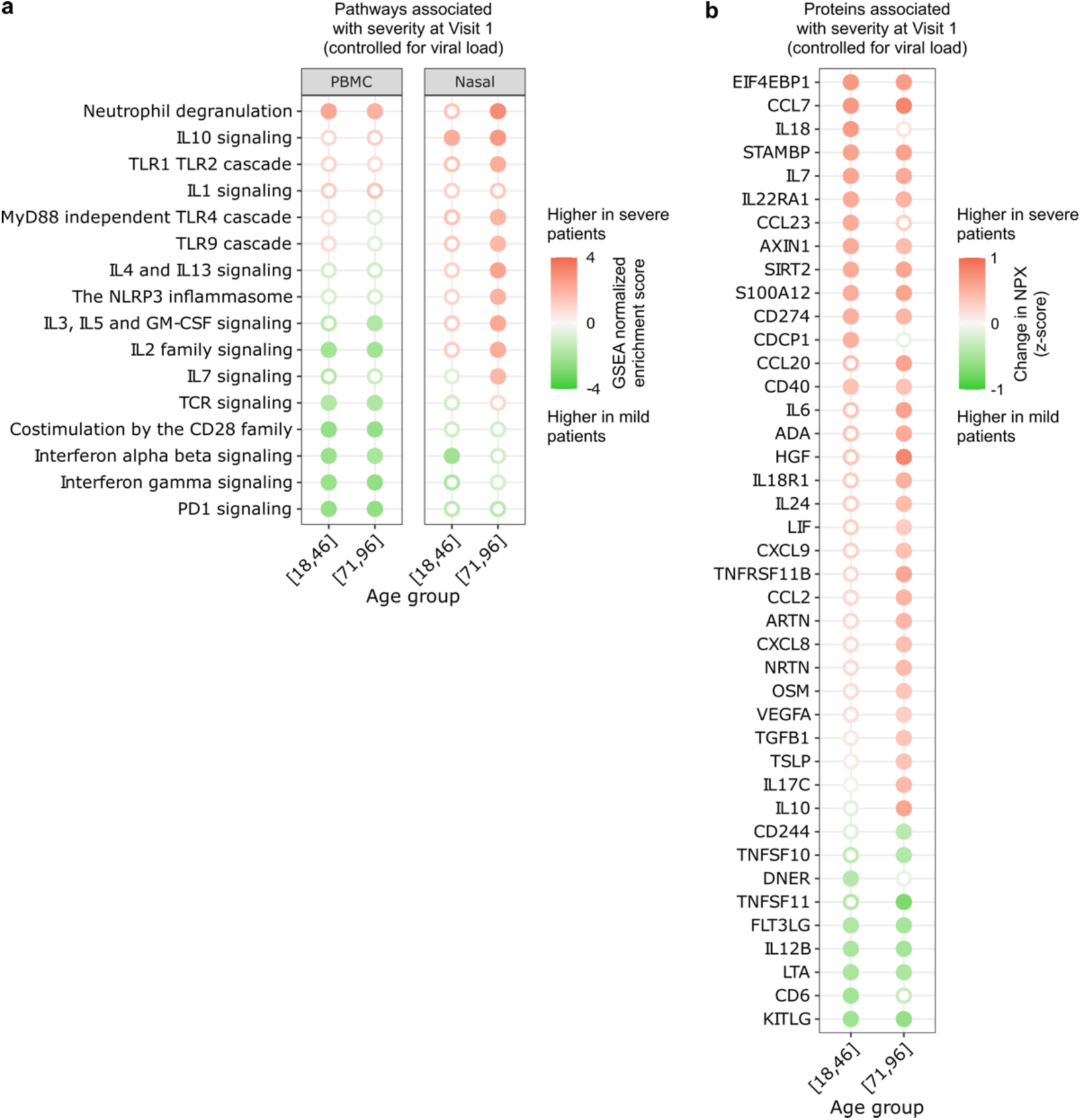
Aging and COVID-19 severity, analyses controlled for viral load. (a, b) Dot plots depicting a) select Reactome pathways in PBMC or nasal RNA-seq data, and b) serum cytokines (olink) that were upregulated in severe patients (NIAID ordinal scales 5-6) compared to mild/moderate (NIAID ordinal scales 3-4) patients at Visit 1, stratified by age group (youngest or oldest). Analogous analyses to Fig. 7, but controlled for viral load.

**Supplementary Figure 10.**
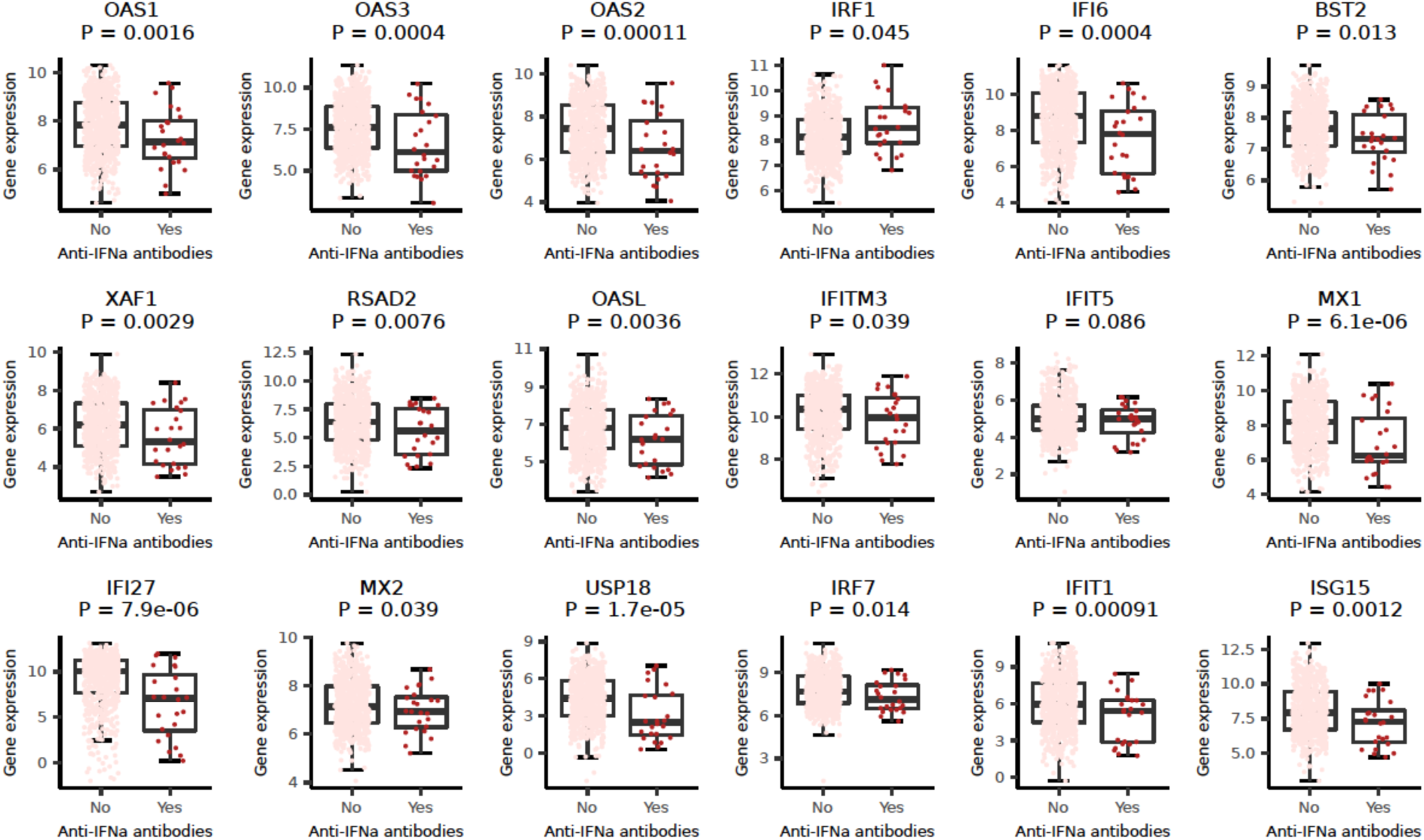
Expression of interferon-related genes in patients with or without anti-IFN-α antibodies at Visit 1. Normalized gene expression is plotted for the subset of samples with both PMBC RNA-seq and anti-IFNα antibody data available (n=732 visit 1 samples). All genes from the Reactome interferon alpha/beta signaling pathway with an adjusted P-value < 0.05 are included (n=18).

**Supplementary Figure 11.**
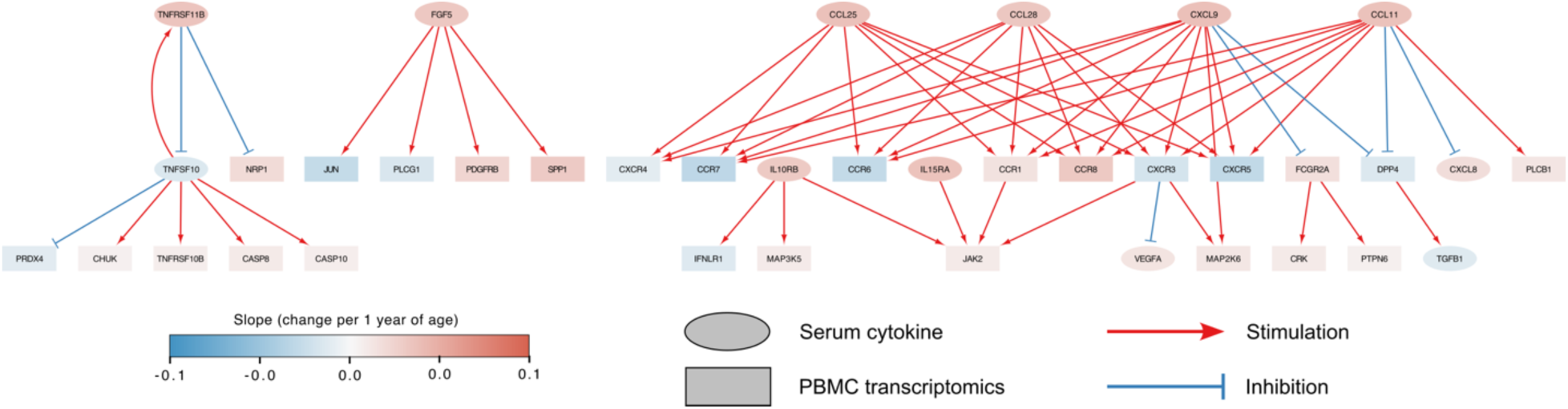
Network analysis of the top 10 significant serum proteins and their receptors and downstream signaling. PBMC RNA-seq and serum cytokine data was integrated using Cytoscape.

**Supplementary Table 1.**
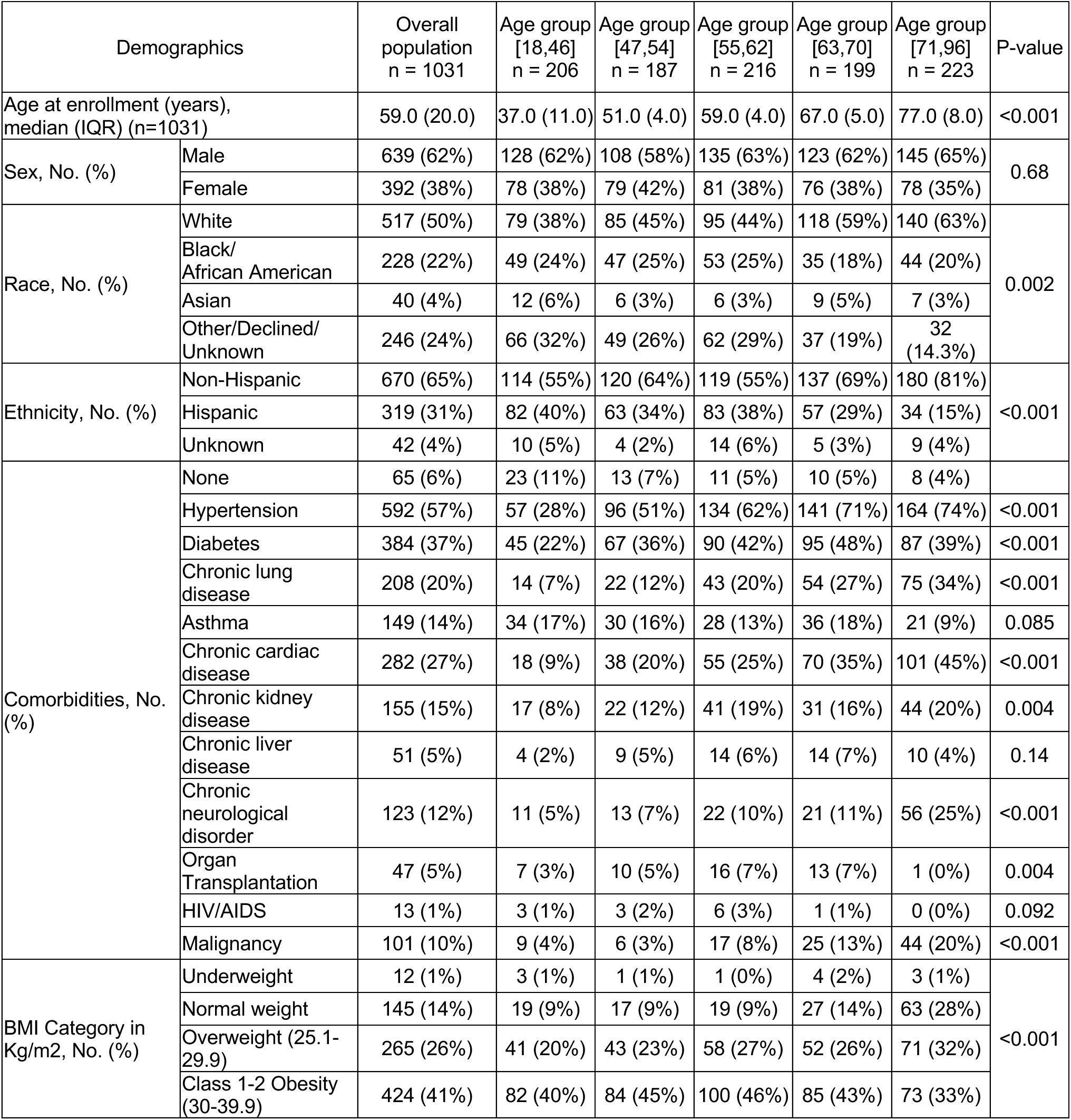
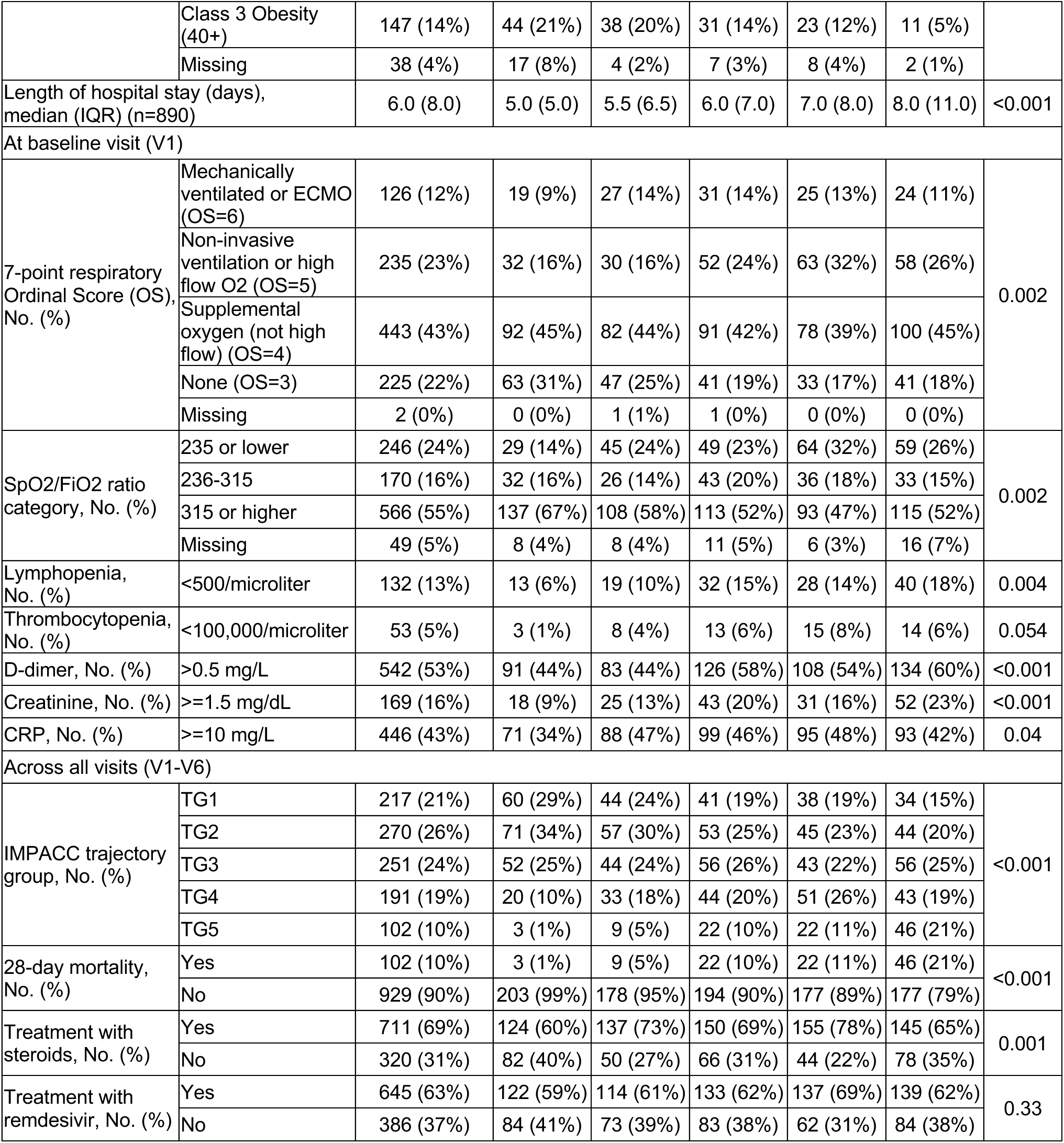
Clinical and demographic characteristics of the cohort at baseline (Visit 1). P values were calculated using Chi-square test for categorical variables, and Kruskal-Wallis test for continuous variables. Percentages might not sum to 100% due to rounding. The number of patients who died within 28 days is the same as the number of patients in trajectory group 5 (TG5).

| Demographics |  | Overall population<br>n = 1031 | Age group<br>[18,46]<br>n = 206 | Age group<br>[47,54]<br>n = 187 | Age group<br>[55,62]<br>n = 216 | Age group<br>[63,70]<br>n = 199 | Age group<br>[71,96]<br>n = 223 | P-value |
| --- | --- | --- | --- | --- | --- | --- | --- | --- |
| Age at enrollment (years), median (IQR) (n=1031) |  | 59.0 (20.0) | 37.0 (11.0) | 51.0 (4.0) | 59.0 (4.0) | 67.0 (5.0) | 77.0 (8.0) | <0.001 |
| Sex, No. (%) | Male | 639 (62%) | 128 (62%) | 108 (58%) | 135 (63%) | 123 (62%) | 145 (65%) | 0.68 |
|  | Female | 392 (38%) | 78 (38%) | 79 (42%) | 81 (38%) | 76 (38%) | 78 (35%) |  |
| Race, No. (%) | White | 517 (50%) | 79 (38%) | 85 (45%) | 95 (44%) | 118 (59%) | 140 (63%) | 0.002 |
|  | Black/<br>African American | 228 (22%) | 49 (24%) | 47 (25%) | 53 (25%) | 35 (18%) | 44 (20%) |  |
|  | Asian | 40 (4%) | 12 (6%) | 6 (3%) | 6 (3%) | 9 (5%) | 7 (3%) |  |
|  | Other/Declined/<br>Unknown | 246 (24%) | 66 (32%) | 49 (26%) | 62 (29%) | 37 (19%) | 32<br>(14.3%) |  |
| Ethnicity, No. (%) | Non-Hispanic | 670 (65%) | 114 (55%) | 120 (64%) | 119 (55%) | 137 (69%) | 180 (81%) | <0.001 |
|  | Hispanic | 319 (31%) | 82 (40%) | 63 (34%) | 83 (38%) | 57 (29%) | 34 (15%) |  |
|  | Unknown | 42 (4%) | 10 (5%) | 4 (2%) | 14 (6%) | 5 (3%) | 9 (4%) |  |
| Comorbidities, No. (%) | None | 65 (6%) | 23 (11%) | 13 (7%) | 11 (5%) | 10 (5%) | 8 (4%) |  |
|  | Hypertension | 592 (57%) | 57 (28%) | 96 (51%) | 134 (62%) | 141 (71%) | 164 (74%) | <0.001 |
|  | Diabetes | 384 (37%) | 45 (22%) | 67 (36%) | 90 (42%) | 95 (48%) | 87 (39%) | <0.001 |
|  | Chronic lung disease | 208 (20%) | 14 (7%) | 22 (12%) | 43 (20%) | 54 (27%) | 75 (34%) | <0.001 |
|  | Asthma | 149 (14%) | 34 (17%) | 30 (16%) | 28 (13%) | 36 (18%) | 21 (9%) | 0.085 |
|  | Chronic cardiac disease | 282 (27%) | 18 (9%) | 38 (20%) | 55 (25%) | 70 (35%) | 101 (45%) | <0.001 |
|  | Chronic kidney disease | 155 (15%) | 17 (8%) | 22 (12%) | 41 (19%) | 31 (16%) | 44 (20%) | 0.004 |
|  | Chronic liver disease | 51 (5%) | 4 (2%) | 9 (5%) | 14 (6%) | 14 (7%) | 10 (4%) | 0.14 |
|  | Chronic neurological disorder | 123 (12%) | 11 (5%) | 13 (7%) | 22 (10%) | 21 (11%) | 56 (25%) | <0.001 |
|  | Organ Transplantation | 47 (5%) | 7 (3%) | 10 (5%) | 16 (7%) | 13 (7%) | 1 (0%) | 0.004 |
|  | HIV/AIDS | 13 (1%) | 3 (1%) | 3 (2%) | 6 (3%) | 1 (1%) | 0 (0%) | 0.092 |
|  | Malignancy | 101 (10%) | 9 (4%) | 6 (3%) | 17 (8%) | 25 (13%) | 44 (20%) | <0.001 |
| BMI Category in Kg/m2, No. (%) | Underweight | 12 (1%) | 3 (1%) | 1 (1%) | 1 (0%) | 4 (2%) | 3 (1%) | <0.001 |
|  | Normal weight | 145 (14%) | 19 (9%) | 17 (9%) | 19 (9%) | 27 (14%) | 63 (28%) |  |
|  | Overweight (25.1-29.9) | 265 (26%) | 41 (20%) | 43 (23%) | 58 (27%) | 52 (26%) | 71 (32%) |  |
|  | Class 1-2 Obesity (30-39.9) | 424 (41%) | 82 (40%) | 84 (45%) | 100 (46%) | 85 (43%) | 73 (33%) |  |
|  | Class 3 Obesity (40+) | 147 (14%) | 44 (21%) | 38 (20%) | 31 (14%) | 23 (12%) | 11 (5%) |  |
|  | Missing | 38 (4%) | 17 (8%) | 4 (2%) | 7 (3%) | 8 (4%) | 2 (1%) |  |
| Length of hospital stay (days), median (IQR) (n=890) |  | 6.0 (8.0) | 5.0 (5.0) | 5.5 (6.5) | 6.0 (7.0) | 7.0 (8.0) | 8.0 (11.0) | <0.001 |
| At baseline visit (V1) |  |  |  |  |  |  |  |  |
| 7-point respiratory Ordinal Score (OS), No. (%) | Mechanically ventilated or ECMO (OS=6) | 126 (12%) | 19 (9%) | 27 (14%) | 31 (14%) | 25 (13%) | 24 (11%) | 0.002 |
|  | Non-invasive ventilation or high flow O2 (OS=5) | 235 (23%) | 32 (16%) | 30 (16%) | 52 (24%) | 63 (32%) | 58 (26%) |  |
|  | Supplemental oxygen (not high flow) (OS=4) | 443 (43%) | 92 (45%) | 82 (44%) | 91 (42%) | 78 (39%) | 100 (45%) |  |
|  | None (OS=3) | 225 (22%) | 63 (31%) | 47 (25%) | 41 (19%) | 33 (17%) | 41 (18%) |  |
|  | Missing | 2 (0%) | 0 (0%) | 1 (1%) | 1 (0%) | 0 (0%) | 0 (0%) |  |
| SpO2/FiO2 ratio category, No. (%) | 235 or lower | 246 (24%) | 29 (14%) | 45 (24%) | 49 (23%) | 64 (32%) | 59 (26%) | 0.002 |
|  | 236-315 | 170 (16%) | 32 (16%) | 26 (14%) | 43 (20%) | 36 (18%) | 33 (15%) |  |
|  | 315 or higher | 566 (55%) | 137 (67%) | 108 (58%) | 113 (52%) | 93 (47%) | 115 (52%) |  |
|  | Missing | 49 (5%) | 8 (4%) | 8 (4%) | 11 (5%) | 6 (3%) | 16 (7%) |  |
| Lymphopenia, No. (%) | <500/microliter | 132 (13%) | 13 (6%) | 19 (10%) | 32 (15%) | 28 (14%) | 40 (18%) | 0.004 |
| Thrombocytopenia, No. (%) | <100,000/microliter | 53 (5%) | 3 (1%) | 8 (4%) | 13 (6%) | 15 (8%) | 14 (6%) | 0.054 |
| D-dimer, No. (%) | >0.5 mg/L | 542 (53%) | 91 (44%) | 83 (44%) | 126 (58%) | 108 (54%) | 134 (60%) | <0.001 |
| Creatinine, No. (%) | >=1.5 mg/dL | 169 (16%) | 18 (9%) | 25 (13%) | 43 (20%) | 31 (16%) | 52 (23%) | <0.001 |
| CRP, No. (%) | >=10 mg/L | 446 (43%) | 71 (34%) | 88 (47%) | 99 (46%) | 95 (48%) | 93 (42%) | 0.04 |
| Across all visits (V1-V6) |  |  |  |  |  |  |  |  |
| IMPACC trajectory group, No. (%) | TG1 | 217 (21%) | 60 (29%) | 44 (24%) | 41 (19%) | 38 (19%) | 34 (15%) | <0.001 |
|  | TG2 | 270 (26%) | 71 (34%) | 57 (30%) | 53 (25%) | 45 (23%) | 44 (20%) |  |
|  | TG3 | 251 (24%) | 52 (25%) | 44 (24%) | 56 (26%) | 43 (22%) | 56 (25%) |  |
|  | TG4 | 191 (19%) | 20 (10%) | 33 (18%) | 44 (20%) | 51 (26%) | 43 (19%) |  |
|  | TG5 | 102 (10%) | 3 (1%) | 9 (5%) | 22 (10%) | 22 (11%) | 46 (21%) |  |
| 28-day mortality, No. (%) | Yes | 102 (10%) | 3 (1%) | 9 (5%) | 22 (10%) | 22 (11%) | 46 (21%) | <0.001 |
|  | No | 929 (90%) | 203 (99%) | 178 (95%) | 194 (90%) | 177 (89%) | 177 (79%) |  |
| Treatment with steroids, No. (%) | Yes | 711 (69%) | 124 (60%) | 137 (73%) | 150 (69%) | 155 (78%) | 145 (65%) | 0.001 |
|  | No | 320 (31%) | 82 (40%) | 50 (27%) | 66 (31%) | 44 (22%) | 78 (35%) |  |
| Treatment with remdesivir, No. (%) | Yes | 645 (63%) | 122 (59%) | 114 (61%) | 133 (62%) | 137 (69%) | 139 (62%) | 0.33 |
|  | No | 386 (37%) | 84 (41%) | 73 (39%) | 83 (38%) | 62 (31%) | 84 (38%) |  |

**Supplementary Table 2.**
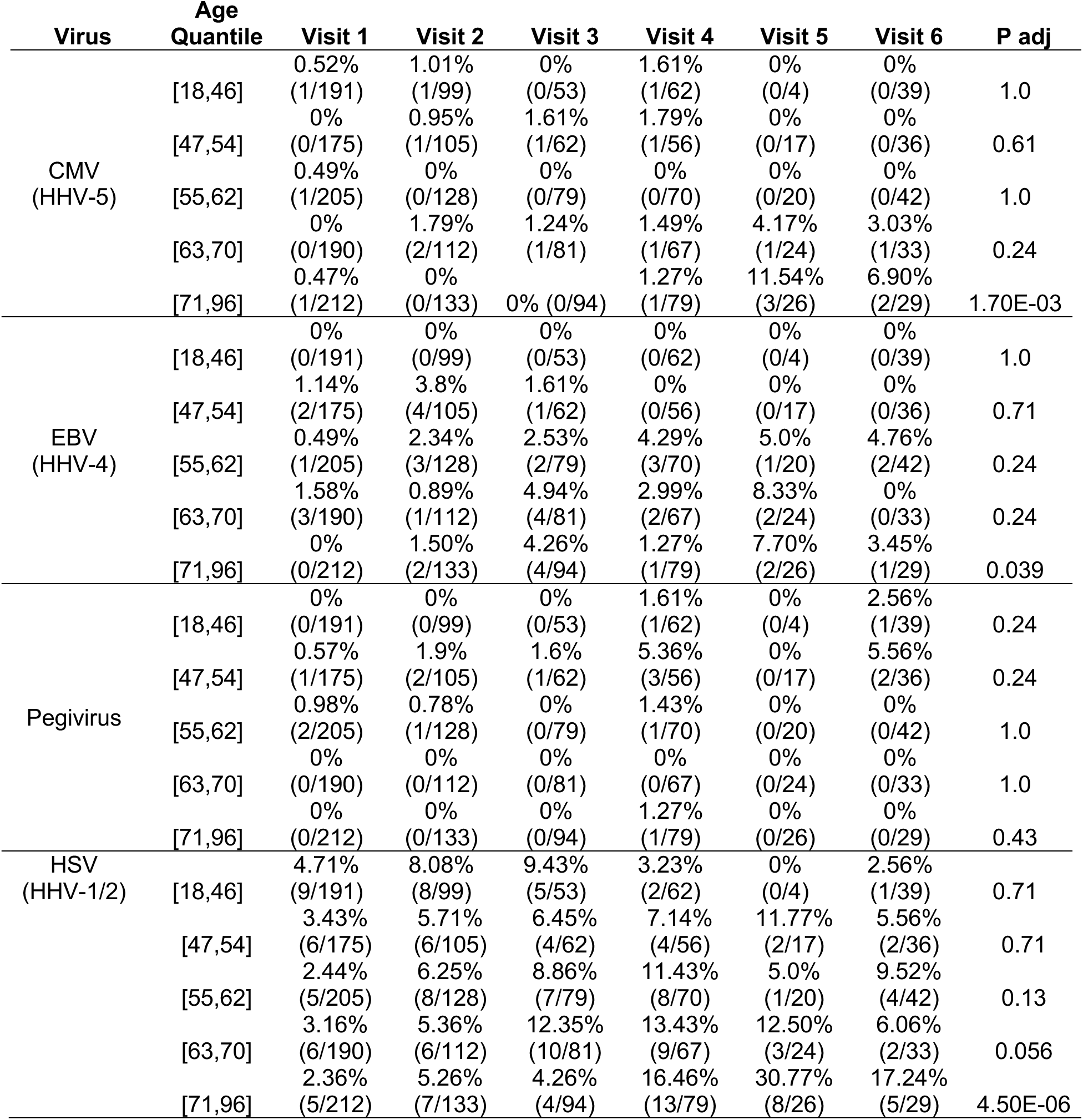
Prevalence of viral cases by age quintile over time in the nasal virome. Adjusted P values (P adj, Benjamini-Hochberg method) are determined by ANOVA with respect to change in prevalence of virus over time within each age quintile. HHV = human herpes virus, CMV = cytomegalovirus, EBV = Epstein Barr virus, HSV = herpes simplex virus.

## Notes

### Competing Interest Statement

The authors have declared no competing interest.

### Author Declarations

NIAID staff conferred with the Department of Health and Human Services Office for Human Research Protections (OHRP) regarding potential applicability of the public health surveillance exception [45CFR46.102(l)(2)] to the IMPACC study protocol. OHRP concurred that the study satisfied criteria for the public health surveillance exception, and the IMPACC study team sent the study protocol, and participant information sheet for review, and assessment to institutional review boards (IRBs) at participating institutions. Twelve institutions elected to conduct the study as public health surveillance, while three sites with prior IRB-approved biobanking protocols elected to integrate and conduct IMPACC under their institutional protocols (University of Texas at Austin, IRB 2020-04-0117; University of California San Francisco, IRB 20-30497; Case Western reserve university, IRB STUDY20200573) with informed consent requirements. Participants enrolled under the public health surveillance exclusion were provided information sheets describing the study, samples to be collected, and plans for data de-identification, and use. Those that requested not to participate after reviewing the information sheet were not enrolled. In addition, participants did not receive compensation for study participation while inpatient, and subsequently were offered compensation during outpatient follow-ups. The International Committee of Medical Journal Editors (ICMJE) recommendations were followed for this study.

